# NeuroPM toolbox: integrating Molecular, Neuroimaging and Clinical data for Characterizing Neuropathological Progression and Individual Therapeutic Needs

**DOI:** 10.1101/2020.09.24.20200964

**Authors:** Yasser Iturria-Medina, Félix Carbonell, Atousa Assadi, Quadri Adewale, Ahmed F. Khan, Robert Baumeister, Lazaro Sanchez-Rodriguez, for the Alzheimer’s Neuroimaging Initiative

## Abstract

There is a critical need for a better multiscale and multifactorial understanding of neurological disorders, covering from genes to neuroimaging to clinical factors and treatments effects. Here we present *NeuroPM-box*, a cross-platform, user-friendly and open-access software for characterizing multiscale and multifactorial brain pathological mechanisms and identifying individual therapeutic needs. The implemented methods have been extensively tested and validated in the neurodegenerative context, but there is not restriction in the kind of disorders that can be analyzed. By using advanced analytic modeling of molecular, neuroimaging and/or cognitive/behavioral data, this framework allows multiple applications, including characterization of: (i) the series of sequential states (e.g. transcriptomic, imaging or clinical alterations) covering decades of disease progression, (ii) intra-brain spreading of pathological factors (e.g. amyloid and tau misfolded proteins), (iii) synergistic interactions between multiple brain biological factors (e.g. direct tau effects on vascular and structural properties), and (iv) biologically-defined patients stratification based on therapeutic needs (i.e. optimum treatments for each patient). All models’ outputs are biologically interpretable. A 4D-viewer allows visualization of spatiotemporal brain (dis)organization. Originally implemented in MATLAB, *NeuroPM-box* is compiled as standalone application for Windows, Linux and Mac environments: neuropm-lab.com/software. In a “regular” workstation, it can analyze over 150 subjects per day, reducing the need for using clusters or High-Performance Computing (HPC) for large-scale datasets. This open-access tool for academic researchers may significantly contribute to a better understanding of complex brain processes and to accelerating the implementation of Precision Medicine (PM) in neurology.

## Introduction

Most prevalent neurological disorders are highly complex, involving a *continuum* of biological alterations from molecular to macroscopic (system level) scales. For instance, Alzheimer’s disease (AD), the most common form of dementia, is characterized by concurrent disruptions in genes, molecular pathways, proteins, vascularity, synapses, neuronal populations, and high-order neuronal- and organ-networks (Palop et al., 2006). It is the continuous crosstalk between these and many other relevant units, as opposed to a unique dominant factor, what causes AD’s associated alterations in memory, thinking, and behavior (Hampel et al., 2019). The massive failing of single-target therapeutic interventions for AD and related disorders, it is clearly evidencing that we cannot really understand, and eventually cure, complex multilevel brain disorders without deeply studying their numerous interrelating components. Furthermore, in keeping with the tenets of Personalized Medicine (PM), clinical treatments require tailoring to multiscale and multifactorial brain mechanisms, and the subsequent individual capacity to response to treatment, contrary to treating all patients with the same approach (Carrasco-Ramiro et al., 2017; Davis et al., 2009; Schork, 2015; Whitcomb, 2012).

In the last decades, the scientific community have gotten much closer to understand the imperative for an integrative (multilevel) analysis of brain’s re-organization and associated diseases. Systems biology, for example, aims to generate spatio-temporal mechanistic models of hierarchical biological networks from normal to pathological conditions (Hampel et al., 2019, 2018). Similarly, the Neuroinformatic field is devoted to the development of analytical and computational models for sharing, integration, and analysis of multimodal neuroscience data (Gaiteri et al., 2018; Mostafavi et al., 2018; Wu et al., 2020; Young et al., 2018; www.incf.org). However, despite the great potential of these disciplines for a better understanding of complex neuropathological processes and the individually-tailored selection of treatments, there is a vital lack of user-friendly open-access tools for both multiscale and multifactorial brain research. This limitation, partly accentuated by the accelerated development of new approaches, contributes to statistical inconsistencies and, traditionally, consumes considerable funding, those being unaffordable for countless experimental groups. Importantly, it is also a major impediment towards fulfilling reproducibility in research.

Motivated by these concerns, we have recently developed a user-friendly software unifying multiple molecular- and neuroimaging-based analytical methods, which have been previously tested and validated in the neurodegenerative context (Iturria-medina et al., 2020; Iturria-Medina et al., 2018, 2017; Yasser Iturria-Medina et al., 2014; Vogel et al., 2020). The *Neuroinformatics for Personalized Medicine* toolbox (*NeuroPM-box*; Fig. 1), constitutes a *long-term* ongoing project aiming to implement, validate and share integrative analytical modeling of molecular, neuroimaging, cognitive/behavioral and/or therapeutic data to better understand individual- or group-level brain (dis)organization mechanisms, as well as identifying personalized therapeutic needs (see neuropm-lab.com/software). Most models’ outputs are biologically interpretable, while a 4D-viewer allows visualization of the brain’s multifactorial spatio-temporal dynamics (e.g. tau and amyloid-β spreading through the cortex). Importantly, there is no restriction in the kind of neuroscience data that it can be applied to. For example, the available methods would be similarly applicable to characterizing multifactorial healthy neurodevelopmental and aging processes. The *NeuroPM-box* will be continuously expanded with new and more integrative methods, with the principal goal of accelerating the understanding of complex brain (dis)organization processes, identifying most relevant disease biomarkers, reducing the cost of patient profiling, and implementing true personalized care in neurology.

**Figure 1.**
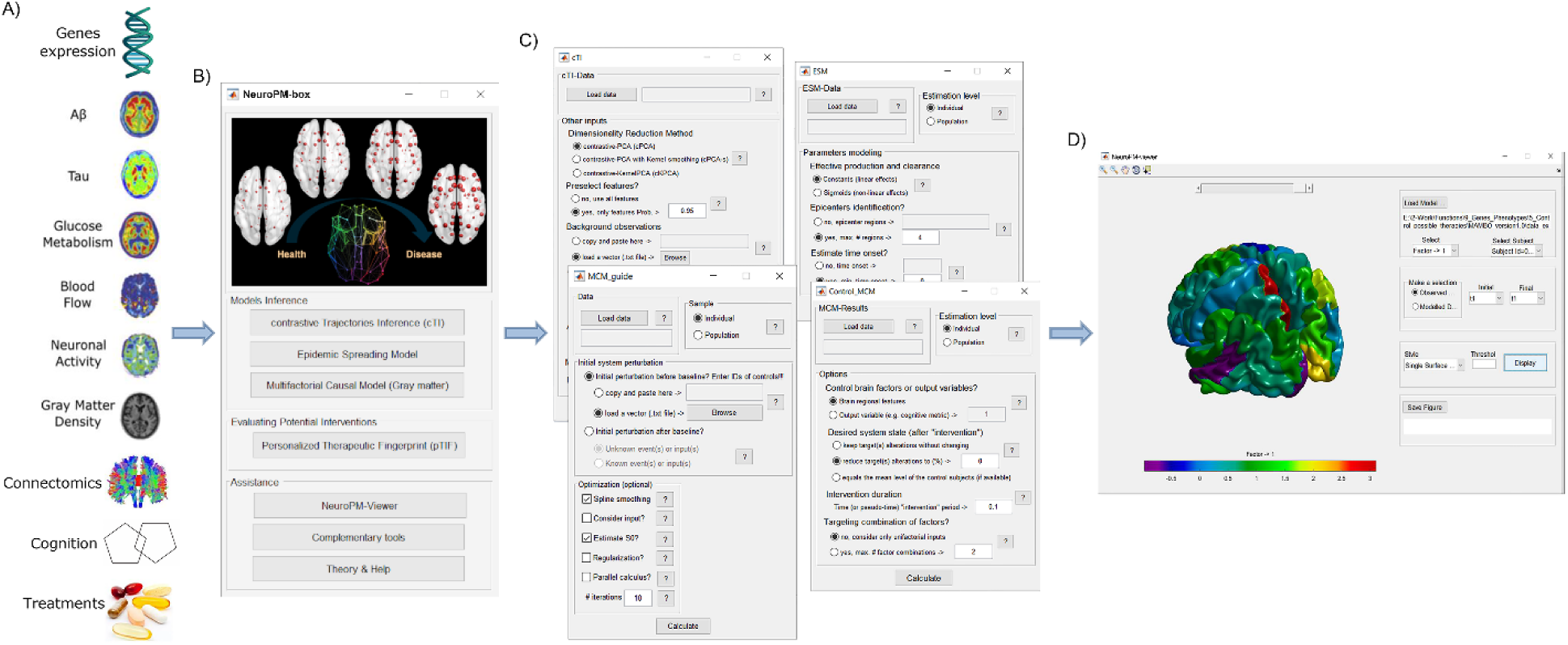
Schematic *NeuroPM-*box software workflow. A) Molecular (e.g. RNA and proteins concentration arrays), multimodal imaging (e.g. tau, amyloid-β and glucose metabolism PET, vascular, functional and structural MRI), whole-brain connectomics (e.g. structural and vascular networks), cognitive/clinical evaluations, and therapeutic interventions data constitute the main expected software inputs. There is not restriction to the number of different modalities that can be analyzed. B) Main *NeuroPM-*box interface, where the user can select from four different analytical methods, apply auxiliary applications, and access the visualization tool and the software’s tutorial. C) Main software modules supporting the data-driven analysis of the multimodal data (available methods are described in next subsections). D) Visualization interface (*NeuroPM-viewer*) allowing detailed exploration over the human cortex of both real and modeled spatiotemporal brain dynamics.

## Software Overview

*NeuroPM-box* (Fig. 1) allows the separate and combined analysis of large-scale molecular and macroscopic data, including molecular screening (transcriptomics, proteomics, epigenomics), histopathology (postmortem neuropathology), molecular imaging (amyloid, tau PET), MRI and cognitive/clinical evaluations. It particularly focuses on clarifying crucial brain mechanistic questions, such as: *i*) which are the series of sequential molecular or macroscopic states (e.g. genetic and brain regional alterations, respectively) underlying decades of neuropathological evolution (Iturria-medina et al., 2020; Young et al., 2014)?; *ii*) which genes (or molecular pathways) drive the dysfunction of other genes and pathways (Iturria-medina et al., 2020; Mostafavi et al., 2018; Zhang et al., 2013)?; *iii*) how disease-agents (e.g. toxic tau and amyloid-β proteins) spread through communicating cells in the brain (Carbonell et al., 2018; Yasser Iturria-Medina et al., 2014; Vogel et al., 2020)?; *iv*) which multifactorial *synergistic* (causal) interactions occur at diseased brain regions (Dyrba et al., 2020; Iturria-Medina et al., 2017)?; and, importantly, *v*) how each patient would respond to different therapeutic interventions (Iturria-Medina et al., 2018)?

The implemented methods and analyses have been developed and successfully validated in multiple previous studies (Iturria-medina et al., 2020; Iturria-Medina et al., 2018, 2017; Yasser Iturria-Medina et al., 2014; Vogel et al., 2020). Crucially, here we aimed to increase the synergy between the different methods and data modalities, allowing to discover new and better ways of multiscale and multifactorial data analysis. Table 1 summarizes the different algorithms and some of the most relevant potential combinations. In brief, the user has access to four main analytical frameworks, their combinations, and a versatile visualization tool, whose performance is illustrated in the following subsections.

**Table 1.**
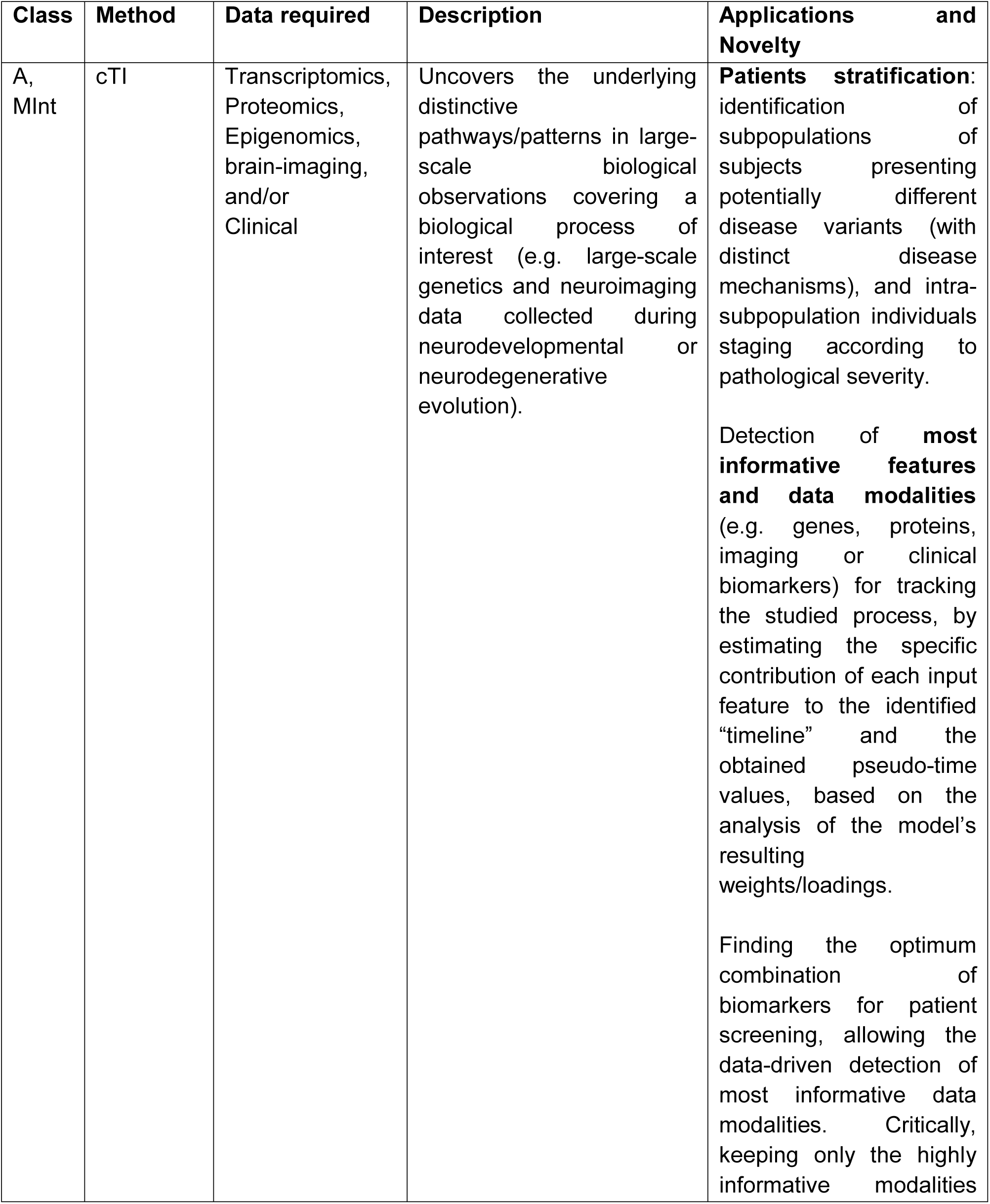
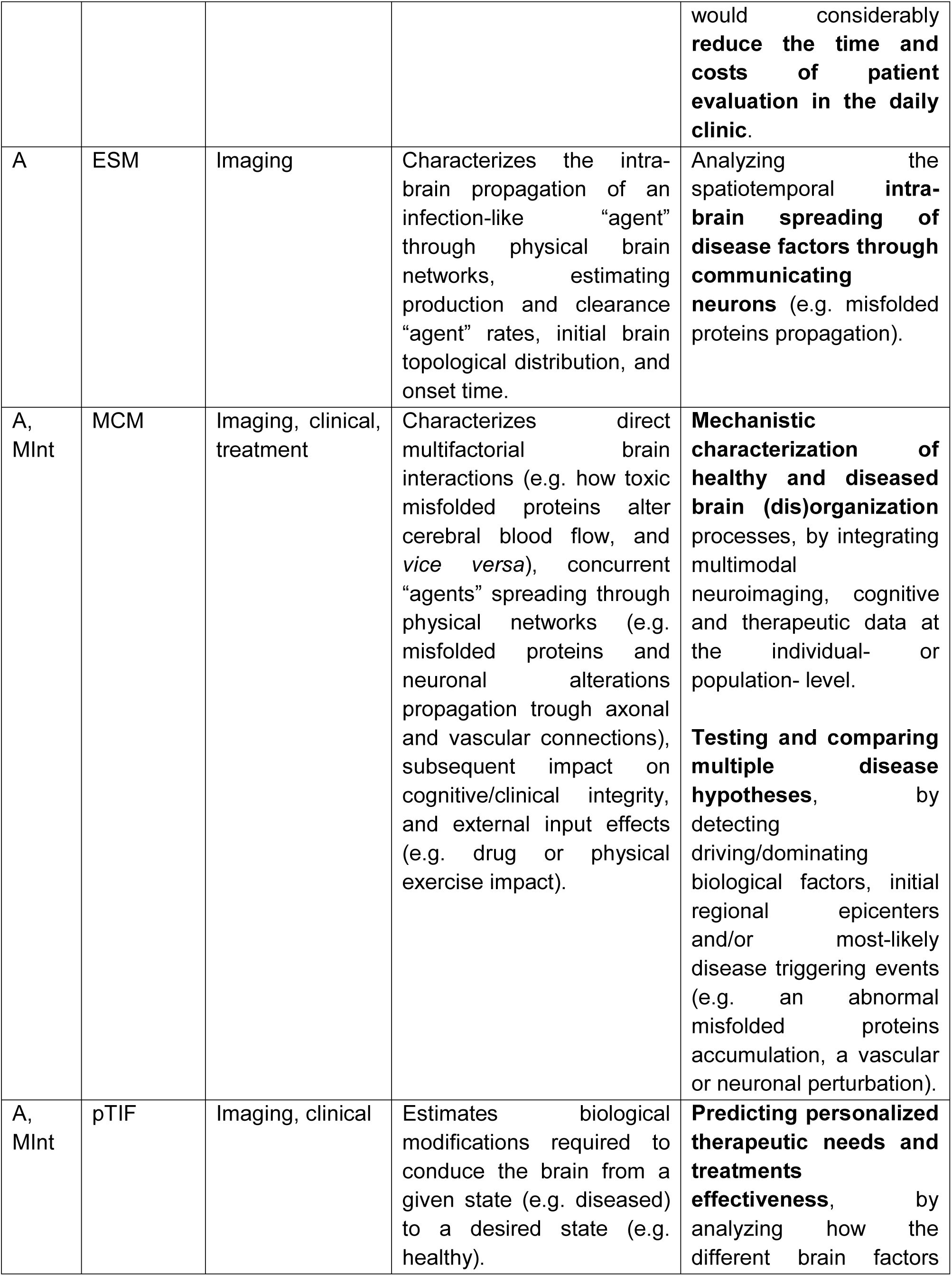
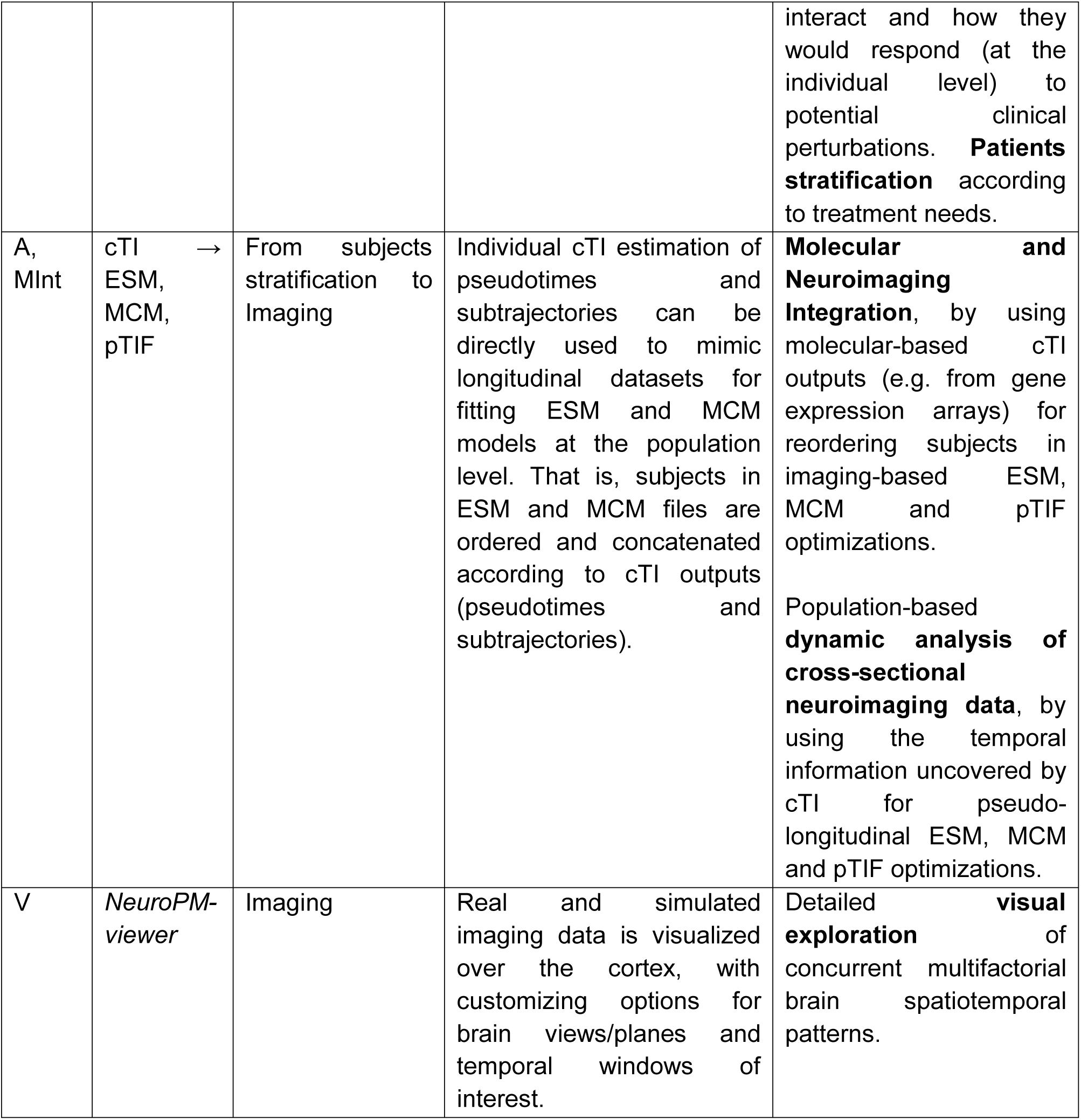
Main *NeuroPM-box* analyses. Each implemented method is available with a detailed tutorial that illustrates how the corresponding applications/modules can be combined for performing complementary analyses. Potential applications are classified as: data Analysis (A), Multimodal Integration (MInt), and data Visualization (V).

| Class | Method | Data required | Description | Applications and Novelty |
| --- | --- | --- | --- | --- |
| A,<br>MInt | cTI | Transcriptomics,<br>Proteomics,<br>Epigenomics,<br>brain-imaging,<br>and/or<br>Clinical | Uncovers the underlying distinctive pathways/patterns in large-scale biological observations covering a biological process of interest (e.g. large-scale genetics and neuroimaging data collected during neurodevelopmental or neurodegenerative evolution). | **Patients stratification:** identification of subpopulations of subjects presenting potentially different disease variants (with distinct disease mechanisms), and intra-subpopulation individuals staging according to pathological severity.<br>Detection of **most informative features and data modalities** (e.g. genes, proteins, imaging or clinical biomarkers) for tracking the studied process, by estimating the specific contribution of each input feature to the identified “timeline” and the obtained pseudo-time values, based on the analysis of the model’s resulting weights/loadings.<br>Finding the optimum combination of biomarkers for patient screening, allowing the data-driven detection of most informative data modalities. Critically, keeping only the highly informative modalities<br>would considerably **reduce the time and costs of patient evaluation in the daily clinic.** |
| A | ESM | Imaging | Characterizes the intra-brain propagation of an infection-like “agent” through physical brain networks, estimating production and clearance “agent” rates, initial brain topological distribution, and onset time. | Analyzing the spatiotemporal **intra-brain spreading of disease factors through communicating neurons** (e.g. misfolded proteins propagation). |
| A, MInt | MCM | Imaging, clinical, treatment | Characterizes direct multifactorial brain interactions (e.g. how toxic misfolded proteins alter cerebral blood flow, and *vice versa*), concurrent “agents” spreading through physical networks (e.g. misfolded proteins and neuronal alterations propagation through axonal and vascular connections), subsequent impact on cognitive/clinical integrity, and external input effects (e.g. drug or physical exercise impact). | **Mechanistic characterization of healthy and diseased brain (dis)organization** processes, by integrating multimodal neuroimaging, cognitive and therapeutic data at the individual- or population- level.<br>**Testing and comparing multiple disease hypotheses,** by detecting driving/dominating biological factors, initial regional epicenters and/or most-likely disease triggering events (e.g. an abnormal misfolded proteins accumulation, a vascular or neuronal perturbation). |
| A, MInt | pTIF | Imaging, clinical | Estimates biological modifications required to conduce the brain from a given state (e.g. diseased) to a desired state (e.g. healthy). | **Predicting personalized therapeutic needs and treatments effectiveness,** by analyzing how the different brain factors interact and how they would respond (at the individual level) to potential clinical perturbations. **Patients stratification** according to treatment needs. |
| A, MInt | cTI → ESM, MCM, pTIF | From subjects stratification to Imaging | Individual cTI estimation of pseudotimes and subtrajectories can be directly used to mimic longitudinal datasets for fitting ESM and MCM models at the population level. That is, subjects in ESM and MCM files are ordered and concatenated according to cTI outputs (pseudotimes and subtrajectories). | **Molecular and Neuroimaging Integration**, by using molecular-based cTI outputs (e.g. from gene expression arrays) for reordering subjects in imaging-based ESM, MCM and pTIF optimizations.<br>Population-based **dynamic analysis of cross-sectional neuroimaging data**, by using the temporal information uncovered by cTI for pseudo-longitudinal ESM, MCM and pTIF optimizations. |
| V | *NeuroPM-viewer* | Imaging | Real and simulated imaging data is visualized over the cortex, with customizing options for brain views/planes and temporal windows of interest. | Detailed **visual exploration** of concurrent multifactorial brain spatiotemporal patterns. |

### Trajectories in large-scale Molecular, Imaging and/or Clinical Data

The *contrasted Trajectories Inference* (cTI) algorithm (Iturria-medina et al., 2020), uses recent AI advances for exploring and visualizing high dimensional data (Abid et al., 2018) with the purpose of uncovering the underlying distinctive/contrasted path in large-scale biological observations (e.g. genetics and neuroimaging data covering a biological process of interest, such as neurodevelopment or neurodegeneration). For instance, in the context of neurodegeneration (see Fig. 2), it allows identifying the series of sequential molecular states (e.g. genetic alterations) covering decades of disease progression, and subsequently revealing the relative position of each individual subject in that path. The algorithm provides a pseudo-time value per subject, reflecting the relative position of each subject on the identified long-term “timeline” (e.g. in neurodegeneration, the pseudo-time have been used as a personalized disease index, significantly predicting individual neuropathology, cognitive deterioration and future clinical conversion). In addition, cTI assigns the subjects to different sub-trajectories in the contrasted space. These sub-trajectories could be reflecting different tendencies in the contrasted data, for example, different disease variants (for a practical example, see results below). Of note, some subjects can have a similar probability to be in multiple sub-trajectories, implying also that sub-trajectories may overlap, particularly at their beginning, where the algorithm may not distinguish between different paths. This could be reflecting real biological effects (e.g. two disease variants with a common or similar starting process). Finally, cTI also allows estimating the specific contribution of each input feature (e.g. gene, imaging biomarker) on the identified “timeline” and the obtained pseudo-time value, based on the analysis of the model’s resulting weights/loadings for each featured included in the original data (i.e. ∼40,000 genes transcripts).

**Figure 2.**
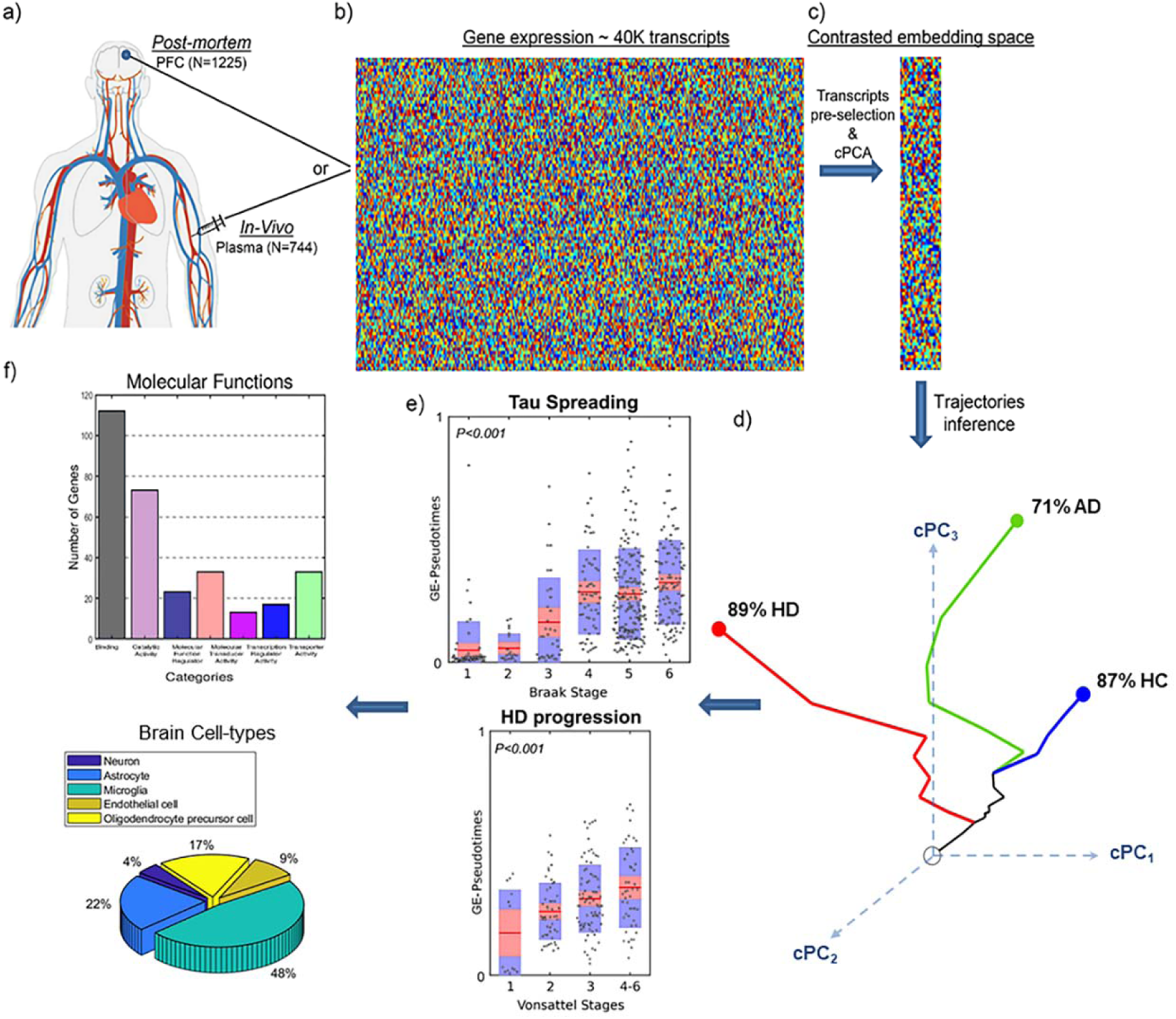
Schematic *cTI* application for detecting disease-associated patterns and patient neuropathological stages in neurodegeneration (Iturria-medina et al., 2020). *In-vivo* blood (N=744; ADNI) and *post-mortem* brain (N=1225; ROSMAP, HBTRC) tissues (**a**) are screened to measure the activity of ∼40,000 transcripts (**b**). Each population’s high dimensional data is reduced to a set of disease-associated components (**c**) via contrastive PCA (Abid et al., 2018; Abid and Zou, 2019). This allow to represent each subject in a disease-associated space (**d**) where the corresponding position reflects her/his pathological state (proximity to the left bottom corner implying a pathology-free state and to the right top corner implying advanced pathology). In this space, each subject is automatically assigned to a disease-trajectory, constituted by the subpopulation of subjects potentially following a common disease variant. The number of subpopulations (disease-trajectories) is automatically determined depending on how the subjects “cluster” together in the disease-associated space. An individual molecular disease score is then calculated, reflecting how advanced each subject is in his/her disease-trajectory. This score significantly predicts neuropathological deterioration (**e**). Finally, the resulting model weights (from contrastive PCA) allow the identification and posterior functional analysis of most influential genes/features (**g**). Panels (a,b,c,e,f) adapted with permission from (Iturria-medina et al., 2020).

Previously, when applied to *in-vivo* microarray gene expression data from the blood of 744 subjects in the Alzheimer’s disease spectrum (ADNI data (Iturria-medina et al., 2020)), cTI automatically identified disease-associated patterns of genes that mirror neuropathological and clinical alterations, and subsequently detects the relative ordering of individuals who better align with those patterns. Similarly, when evaluated on 1225 post-mortem brains in the spectrum of AD and Huntington’s disease (HD; ROSMAP and Harvard Brain Tissue Resource Center data [HBTRC]), it strongly predicted neuropathological severity and comorbidity (Braak, Amyloid and Vonsattel stages; see Fig. 2d-e for results in HBTRC). In consistence with findings in ADNI and ROSMAP (Iturria-medina et al., 2020), in the HBTRC data, we observed (Figs. 2e) a positive association between the individual molecular disease score and the levels of neuropathologic affectation in AD and disorders Huntington’s disease (HD). The GE-pseudotimes were significantly associated with the Braak stages (Fig. 2e (top); F=11.17, P<0.001, FWE-corrected) and the Vonsattel stages (Fig. 2e (bottom); F= 9.04, P<0.001, FWE-corrected).

Here, we also explored cTI’s capacity to distinguish neurological conditions in a highly heterogeneous population. For this, we separately reanalyzed HBTRC’s GE and histopathological data, including two different disorders (LOAD and HD) and nondemented controls (Dataset 1, N=736; see *Materials* and *Procedure*). Using each data modality (GE or histopathology), the method automatically identified multiple sub-trajectories reflecting diagnosis-specific subpopulations (Fig. 2d). For instance, based on GE, the identified sub-trajectory 1 was constituted by 87% of the nondemented controls, 18% of the AD and 2.7% of the HD subjects; a sub-trajectory 2 presented 71% of the AD, 38% of the controls and 12% of the HD participants; and a sub-trajectory 3 was constituted by 89% of the HD, 32% of the nondemented, and 25% of the AD subjects. Similarly, based on a limited set of histopathological data (just 25 broad atrophy metrics; see *Example Dataset 1*), the sub-trajectory 1 presented 91% of the nondemented controls, 28% of AD and 4.9% of HD; a sub-trajectory had 62% of the HD, 57% of the AD and 40% of the controls; and a last sub-trajectory 3 was constituted by 48 of the HD, 26% of the AD and 2.3% of the controls. All together, these results support that cTI is a promising technique for patient stratification in terms of disease stages and variants, even in the presence of comorbid neurological conditions.

In addition to identifying individual disease stages and variants, and corresponding most informative features, cTI offers the possibility to extract the intrinsic dynamic information in a large-scale cross-sectional data. As described in Table 1 (Application *cTI* → *ESM, MCM, pTIF*), this functionality can be particularly useful for analyzing cross-sectional neuroimaging studies as if they were longitudinal. That is, individual pseudotimes and subtrajectories can be used to mimic longitudinal datasets for fitting ESM, MCM and pTIF models at the subpopulations level. Specifically, using the “Adding new stratification to ESM/MCM files” option in “Complementary Tools”, subjects in ESM and MCM files can be ordered and concatenated according to cTI outputs (pseudotimes and subtrajectories) or according to any user provided patients stratification, based on clinical information or obtained via another computational method (Park et al., 2017; Young et al., 2018).

### Epidemic Spreading of Pathological Agents

The epidemic spreading model (ESM) in the neurological context (Yasser Iturria-Medina et al., 2014) characterizes the intra-brain propagation of infection-like “agents” through physical brain networks (e.g. anatomical, vascular, functional). The model (Fig. 3) estimates individual rates of “agent” clearance and production, which can follow linear or sigmoid relationships with the local concentration of the modeled infection-like “agents”. ESM has been successfully applied to further understand amyloid and tau misfolded proteins spreading in the human neurodegenerative brain (Yasser Iturria-Medina et al., 2014; Vogel et al., 2020).

**Figure 3.**
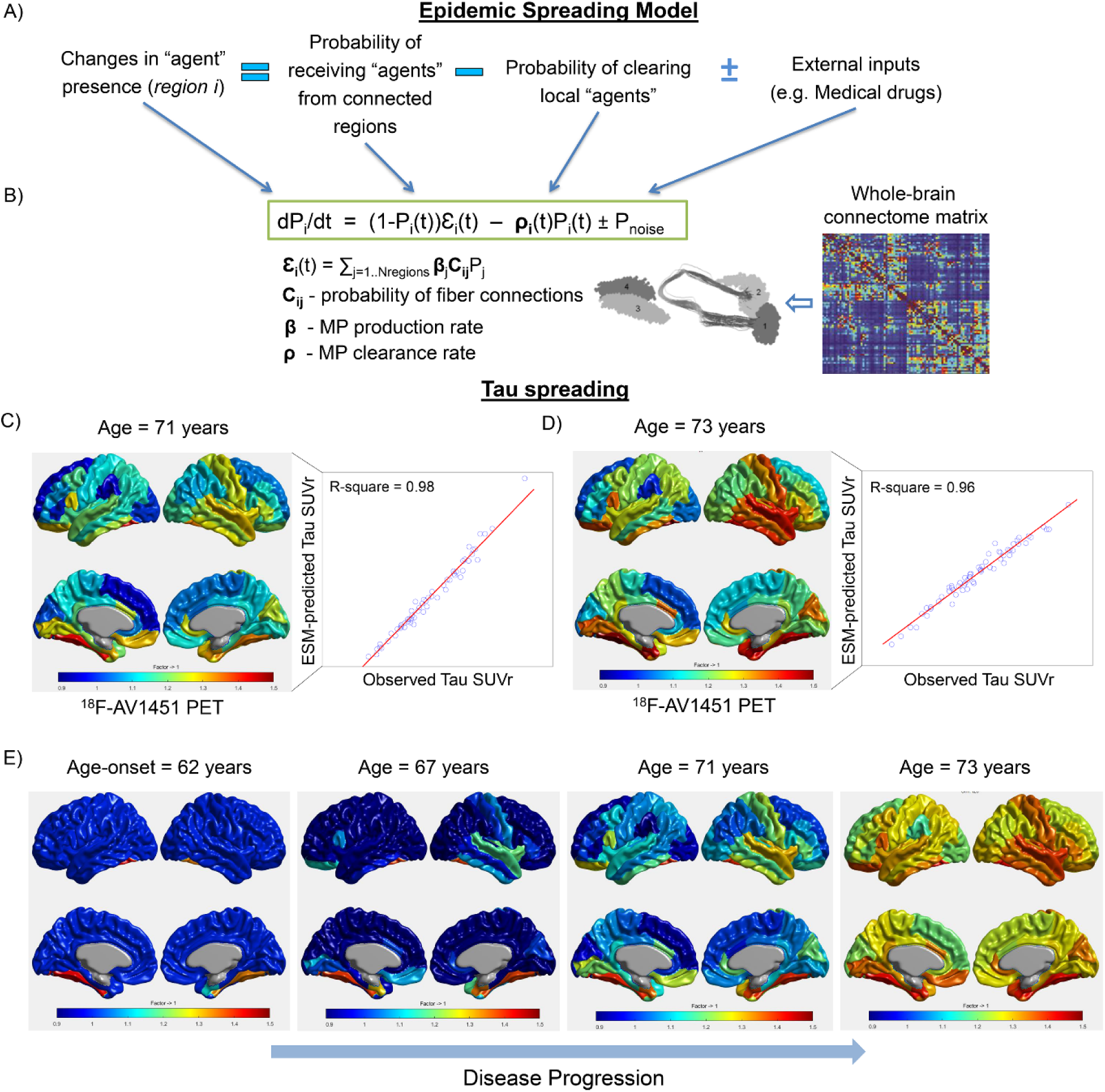
ESM approach and results on tau PET. A) Changes in the presence of a given infection-like “agent” factor (e.g. amyloid and tau misfolded proteins) at an specific brain region *i* are modeled as a function of the incoming “agents” from each connected region *j* (i.e. spreading effects through communicating cells) minus the local “agent” clearance, plus the effects of external inputs (e.g. treatments, physical exercise). B) This cause-effect dynamic model can be mathematically translated to a non-linear system of differential equations, which is dependent on the individual “agent” production rate, the clearance rate, and the inter-regions brain connectivity matrix. In the *NeuroPM-box*, both the production and the clearance rates can be optionally modeled as time-dependent sigmoid functions (Yasser Iturria-Medina et al., 2014) or as global constant values. The connectivity matrix can be estimated via diffusion MRI tractography (Hagmann et al., 2007; Iturria-Medina et al., 2007) or an alternative technique (Bakker et al., 2012; Friston et al., 2014a; Wig et al., 2011). C) ESM results reproducing the tau deposition patterns at the first ^18^F-AV1451 PET evaluation (age=71 years) of a female clinically healthy control with significant memory complaints (ADNI data, subject ID 024_S_5290). D) ESM results in the same participant at the fourth time point evaluation (age=73 years). Starting from a pathology-free stage, the model explained an 86% (P<10^−10^) of the variance in tau values across the four available time points. E) ESM simulation of the whole-brain intra-brain tau spreading process from the estimated onset-time of tau appearance/propagation (age=62 years) to the last observed time point. In C-E, tau values are cortical-to-cerebellum Standardized Uptake Value ratio (SUVr).

Importantly, the current model implementation presents significant improvements compared to the initial version applied in (Yasser Iturria-Medina et al., 2014; Vogel et al., 2020). The three main enhancements are: i) focusing on individual longitudinal data, allowing to fit as many time points as available and subsequently increasing the model parameters’ robustness and biological interpretably, ii) working with the original imaging signal (e.g. SUVr values from PET) or with probabilistic-inferred values from the original images (the original model was defined only for probabilistic values), and iii) covering all the possible numeric values for parameters optimization, the differential equations are solved now with a considerably more robust algorithm, improving the finding of global solutions (instead of potential local minimums) and, subsequently, increasing parameters robustness and interpretably.

Here, the ESM was applied to a population of 105 healthy and diseased subjects with at least two longitudinal tau PET acquisitions (^18^F-AV1451 ligand, ADNI data; see *Materials* and *Procedure*). In average, when applied at the individual level, this approach explained the 80% (std 9.6) of the variance in regional tau values across all available time points. Figure 3 shows the model’s results in a female clinically healthy control participant with significant memory complaints and four longitudinal tau PET evaluations (ADNI data, subject ID 024_S_5290). Starting from a pathology-free stage, the model explained a 86% (P<10^−10^) of the variance in tau values across the four available time points (Figs. 3C,D). Notice the strong correspondence between the observed and reproduced tau deposition patterns at the first and last PET evaluations, ages 71 (Fig. 3C right) and 73 (Fig. 3D right), respectively. In addition, it automatically identified the entorhinal cortex, fusiform gyrus, caudate anterior cingulate and inferior parietal in the left hemisphere as most-likely tau spreading epicenters in this subject and estimated that tau accumulation and spreading started about the year 62 of her life. Figure 3E illustrates the long-term intra-brain propagation process, starting on the identified epicenters and diffusing for over a decade to the other brain regions, following a stereotypic AD-related pattern (Braak and Braak, 1991). In this analysis, we allowed up to a maximum of four regions as initial epicenters and a maximum value of 0.1959 (i.e. 5% of maximum observed SUVr) for non-epicenter regions at the onset time.

### Multifactorial causal Model of Brain (Dis)organization

The multifactorial causal model of brain (dis)organization and cognition (MCM (Iturria-Medina et al., 2017)), accounts for: i) regional multifactorial causal interactions (e.g. how toxic misfolded proteins alter cerebral blood flow [CBF], how subsequent alterations in CBF influences neuronal activity and gray matter atrophy, and *vice versa*), ii) concurrent perturbations propagation through physical networks (e.g. intra-brain propagation of misfolded proteins, vascular or neuronal alterations across axonal and vascular connectomes), iii) the subsequent impact of (i) and (ii) on cognitive/clinical integrity, and iv) global factor-specific effects of external inputs (e.g. how a given clinical treatment impacts tau, amyloid and CBF). The MCM considers that once a factor-specific event occurs in a given brain region or in a set of regions, it can directly interact with other biological factors and alter their states. The alterations can also spread through physical connections (e.g. anatomical, vascular connections) to other brain areas, where similar factor-to-factor and propagation mechanisms may occur, in a continuous cycle. The MCM has been successfully applied to the study of Alzheimer’s disease (Iturria-Medina et al., 2018, 2017), clarifying multifactorial disease-specific mechanisms, and is currently being applied to other neurodegenerative disorders (e.g. Amyotrophic lateral sclerosis, Frontotemporal dementia, Parkinson’s disease).

The currently implemented version presents multiple key improvements with regards to the initial model version (Iturria-Medina et al., 2017). The two main enhancements are: i) focusing on individual longitudinal data (in addition to allow group level fitting as in the initial article), allowing to fit as many individual time points as available and increasing the model parameters’ biological interpretably, and ii) covering all the possible numeric values for parameters optimization, the differential equations are solved now in a considerably more robust way, improving the finding of global solutions (instead of potential local minimums) and, subsequently, increasing parameters robustness and interpretably.

Here, the MCM approach was applied to a healthy and diseased population of 504 participants (ADNI data, see *Materials* and *Procedure*) with 4 to 6 different imaging modalities and at least 4 longitudinal evaluations. The imaging modalities included tau-PET, amyloid-PET, FDG-PET (for quantifying glucose metabolism), resting-fMRI (for neuronal activity at rest), ALS-MRI (for cerebral blood flow) and structural-MRI (for gray matter density). For all subjects, the average longitudinal time window was 5.1 years (std=3.6). The model’s numerical optimization converged successfully in 496 (98.4%) subjects of the population. Starting from a free-pathology state, the MCM explained in average 92% (std=4.7) and 71% (std=15.1) of the first and last individual multimodal observations, respectively. Figure 4C illustrates the MCM-simulated concurrent intra-brain changes in tau, amyloid and gray matter atrophy on a female clinically healthy control with significant memory complaints (ADNI data, subject ID 024_S_5290). Notice that this is the same participant previously analyzed with ESM [Fig. 3C-E], but now characterized from a multifactorial perspective (i.e. going from a univariate tau analysis to an integrative multimodal characterization). In this subject, for the first (67 years) and last observed time points (73 years), the model explained 94.6% and 51.9% of the variance across the six data modalities, respectively. Data simulation (Fig. 4C) is extended for three additional years after the last time point (i.e. from 73 to 76 years), illustrating the model’s capacity to predict future disease progression. In this specific case (Fig. 4C), notice a prominent increase in tau brain deposition in parallel to a substantial reduction of the brain’s gray matter density. In addition, for each individual, MCM is able to characterize the factors-specific tendencies to spread through the anatomical connectome, i.e. how alterations in each considered biological factor (e.g. tau, amyloid) propagate from region to region (data not shown). Factor-to-factor direct interactions, typically reflecting a strong synergy between multiple relevant components in AD progression, are also characterized (Iturria-Medina et al., 2017). Other interesting MCM outputs include the estimation of the first pathological perturbation in the disease process (i.e. the brain regions and biological factors that are initially altered), the specific influence that each biological factor has on the other factors, the proportion of each factor’s longitudinal changes caused by other factors and by itself, and, if applicable, the global (across regions) impact that a given treatment has on each biological factor (see software Tutorial).

**Figure 4.**
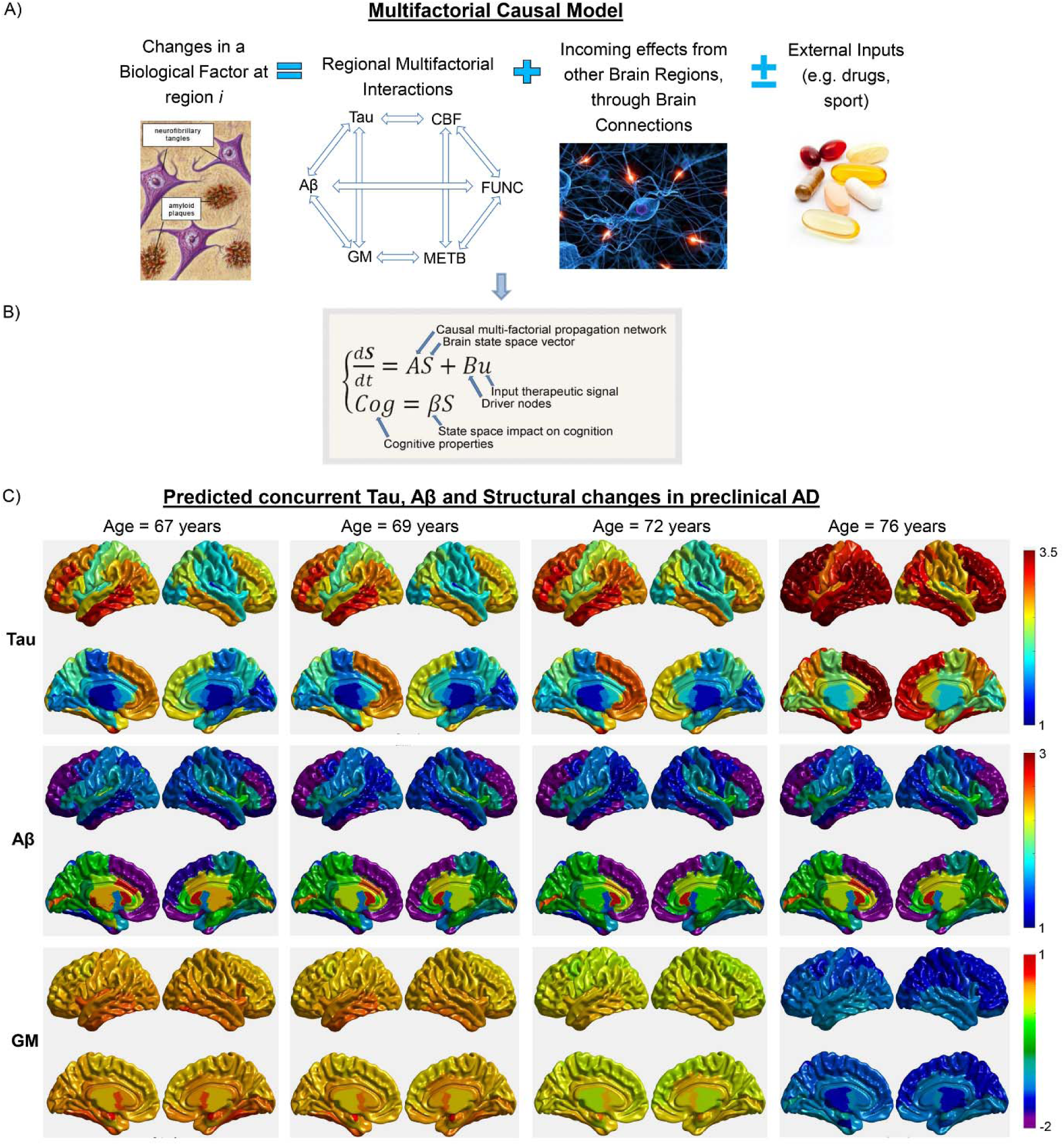
MCM approach and results on six different imaging modalities. A) Changes in a given biological factor (e.g. amyloid and tau deposition) at an specific brain region are modeled as a function of the local multifactorial synergistic interactions (e.g. how cerebrovascular flow dysregulation influence amyloid and tau deposition), the intra-brain alterations spreading through communicating cells, and external inputs (e.g. treatments). B) This cause-effect dynamic model can be mathematically translated into a system of differential equations. Similarly to previously proposed brain causal models (Friston et al., 2014b; Y Iturria-Medina et al., 2014; Stephan et al., 2010; Valdes-sosa et al., 2011), in MCM causality is intrinsic in its differential equations. But, going beyond the traditional single-factor modeling (commonly neuronal activity or misfolded proteins), the MCM equations describe: (i) how the present state of a given biological factor, at a given brain region, causes new fluctuations in other factors or itself at the same region or at a different brain region, via multifactorial local interactions or by spreading through brain connections, and (ii) how the system’s dynamic may change due to the influence of external inputs (e.g. cognitive/sensory stimulus, therapeutic interventions or environmental influences). C) MCM-simulated concurrent intra-brain changes in tau, amyloid and gray matter (GM) density on a female clinically healthy participant with significant memory complaints (ADNI data, subject ID 024_S_5290). The simulation is based on the model parameters obtained for this subject, while an additional time window (from 73 to 76 years) of multifactorial data was generated for prediction purposes. Notice a prominent increase in tau brain deposition in parallel to a substantial reduction of the brain’s gray matter density. In (A), FUNC refers to functional activity at rest (e.g. from fMRI), and METB to glucose metabolism (e.g. from FDG PET).

### Imaging-based therapeutic fingerprints

The *personalized Therapeutic Intervention Fingerprint* (pTIF; Iturria-Medina et al., 2018, *Neuroimage*) assumes that the patients in a heterogeneous population need different treatments, not only depending on their brain’s unifactorial alterations (e.g. tau/amyloid deposition or not, cerebrovascular alterations or not, atrophy or not) but also on their individual multifactorial brain dynamics: how the different biological factors interact and how they would respond (at the individual level) to potential clinical perturbations. Based on spatiotemporal analysis of multi-modal imaging data (i.e., PET, MRI, SPECT), pTIF values are a set of multivariate metrics that reflect the biological changes required to stop a given brain reorganization process or revert the condition to normality. In other words, the pTIF can integrate large amount of data (e.g. thousands of multi-modal brain imaging measurements) into a simplified individual patient profile -the *fingerprint*-of the quantitative biological factor modifications needed to control the reorganization process (e.g. disease evolution). Results in aging and late-AD (ADNI data) support that pTIF allows to categorize the patients into distinctive therapy-based subtypes, which are in strong correspondence with differential genetic and cognitive profiles.

### Visualizing Observed and Modelled Spatiotemporal Brain Dynamics

Visualization of both acquired and simulated brain data is crucial for a deeper understanding of the studied brain processes. *NeuroPM-box* include a versatile user-friendly interface which allows visualizing all analyzed brain factors and their dynamic changes, in a single, 4 or 8 surfaces view. The user can show observed or simulated data at the individual- or group-level. Time is one of the most important variables for both modeling and visualization, subsequently, this viewer allows handling the time variable according to the specific visualization needs. Other settings (colormaps, minimum and maximum values, threshold) can be manually adjusted. Finally, this tool allows saving the depicted figures in traditional images formats (.jpg, .png, and .tif) and dynamic brain videos (.avi).

### Required Expertise

The *NeuroPM-box* is deliberately designed to be a postprocessing analytic software and not a neuroimaging preprocessing package, for which many excellent open-access software already exist (e.g. SPM, FSL, ANTS, CIVET, FreeSurfer, MRtrix3, DSI studio, BrainSuite). Consequently, when using the imaging-based ESM, MCM and pTIF approaches, all basic imaging preprocessing (i.e. across-modalities registration to a unique space, brain parcellation, connectomes estimation) should be previously performed by a user with neuroimaging preprocessing expertise. Similarly, any preprocessing step required for molecular data analysis (e.g. transcriptomics, epigenomics and proteomics quality control) should be previously performed. In addition, the user should have basic expertise writing/reading numerical data in text files, and, when using a large-scale population, should be able to organize the data in the required format (see *Materials and Procedure*), ideally using automatic scripts. Importantly, advanced mathematical and/or computational knowledge is not required, since all the models can be understood in intuitive biological terms.

### Comparison with other software

To the best of our knowledge, at present no other across-platform open-access user-friendly software exists for integrating large-scale molecular, macroscopic and clinical data via advanced mathematical modeling. *NeuroPM-box* allows separated and combined analysis of molecular screening (transcriptomics, proteomics, epigenomics), histopathology (postmortem neuropathology), molecular imaging (amyloid, tau PET), macroscopic MRI and cognitive/clinical evaluations. Most available packages focus uniquely on either molecular (Bézieux et al., 2020; Bosco et al., 2019; Trapnell et al., 2014) or imaging analysis, and not on their possible combination, which *NeuroPM-box* is designed for. In addition, models for characterizing intra-brain spreading of brain alteration effects (e.g. ESM, MCM) or for identifying individual therapeutic needs (e.g. pTIF) had not been added to any other user-friendly software. Similarly, some computational codes for patient stratification in the neurological context have been shared (Park et al., 2017; Young et al., 2018), but their application requires user programming skills.

Importantly, due to the recentness of all the implemented methods (Iturria-medina et al., 2020; Iturria-Medina et al., 2018, 2017; Yasser Iturria-Medina et al., 2014), we believe that their user-friendly implementation will accelerate the comparison, and even the potential integration, with many different methods recently proposed by other teams (Park et al., 2017; Raj et al., 2012; Trapnell et al., 2014; Young et al., 2018). In this sense, the novelty of the research questions tackled by ours and other groups’ novel approaches, strongly supports the crucial need for further developing and extending open-access user-friendly software packages like *NeuroPM-box*.

### Limitations and Future work

As mentioned, *NeuroPM-box* is a *long-term* ongoing project implementing, validating and sharing integrative analytic brain modeling for better understanding of brain (re)organization processes and treatments selection (neuropm-lab.com/software). All presented methods are subjected to a continuous revision, particularly for improving their numerical optimization (an open-ended field in research) and their results interpretation/visualization. New methods are currently under development, with the main purpose of further integrating multiscale and multimodal neuroscience research. Subsequently, many methodological modifications will be implemented in the coming years. By sharing *NeuroPM-box*, we hope to receive continuous feedback on the identification of remaining limitations and translational gaps, while accelerating the clinical use and further validation of the developed methods and, consequently, increasing their worldwide applicability.

For the appropriate use of *NeuroPM-box*, multiple factors should be carefully considered while some challenges remain. As discussed above, the software is not intended to be a neuroimaging preprocessing package. Therefore, when using neuroimaging data, the user should provide the quality-controlled data organized into a specific format (see Tutorial), with all intra-individual images registered to a same native or normalized space (i.e. same images orientation, dimensions and resolution). Future software versions will focus on softening this condition, by allowing to read different imaging modalities written at different spatial dimensions and resolution. For facilitating quality control, specific outlier detection methods are currently implemented (e.g. using the three sigma rule) as well as data correction and completion via imputation (Folch-Fortuny et al., 2016). Also, in order to keep increasing the software’s generalizability, we are planning to extend it to input data from popular organization formats, including the *Brain Imaging Data Structure* (BIDS) standard (Gorgolewski et al., 2016).

Current *NeuroPM-box* data visualization module is restricted to the brain’s cortical surface. This hinders the observation of subcortical patterns. In addition, surface visualization is strongly dependent on the used brain parcellation. For instance, presenting interpolation and signal contamination issues for volumetric brain parcellations for which the cortical surface mesh is not available. To overcome these limitations, we are currently designing and developing a complementary 3D volume-based visualization interface. In the near future, the user will be able to plot the observed and simulated data both on the cortical surface and the whole brain volume.

A common concern when applying advanced neuroscience computational techniques is the running time. Most of the *NeuroPM-box*’s optimization algorithms have been implemented to minimize computational time. For instance, cTI can analyze thousands of subjects and large-scale omics data in just a few minutes. However, the differential equations-based methods (ESM, MCM and pTIF) are more computationally expensive, particularly when applied at the individual level. The implemented ‘default’ optimization for these methods can significantly reduce the computational time, without compromising the accuracy of the solution, in comparison to other available techniques (i.e. MATLAB’s trust-region-reflective algorithm (Coleman and Li, 1996)). Yet, when analyzing over hundreds of subjects with multimodal imaging and longitudinal data, a regular workstation could take from a few days to a week (depending on the number of imaging modalities, brain regions and time points). Because of this, we are currently evaluating to upload the software to popular High-Performance Computing (HPC) portals, for example, The Neuroscience Gateway (NSG, http://www.nsgportal.org) and CBRAIN (http://www.cbrain.ca/).

## Materials

### Example Dataset 1

736 individual *post-mortem* tissue samples from the dorsolateral prefrontal cortex BA9 of LOAD patients (N=376), HD patients (N=184) and nondemented subjects (N=173) were collected and analyzed (Zhang et al., 2013). All autopsied brains were collected by the Harvard Brain Tissue Resource Center (HBTRC; GEO accession number GSE44772), and include subjects for whom both the donor and the next of kin had completed the HBTRC informed consent (http://www.brainbank.mclean.org/). Correspondingly, tissue collection and the research were conducted according to the HBTRC guidelines (http://www.brainbank.mclean.org/). Postmortem interval (PMI) was 17.8 ± 8.3 hr, sample pH was 6.4 ± 0.3 and RNA integrity number (RIN) was 6.8 ± 0.8 for the average sample in the overall cohort. As described in (Zhang et al., 2013), RNA preparation and array hybridizations applied custom microarrays manufactured by Agilent Technologies consisting of 4,720 control probes and 39,579 probes targeting transcripts representing 25,242 known and 14,337 predicted genes. Arrays were quantified on the basis of spot intensity relative to background, adjusted for experimental variation between arrays using average intensity over multiple channels, and fitted to an error model to determine significance (Emilsson et al., 2008). Braak stage, general and regional atrophy, gray and white matter atrophy and ventricular enlargement were assessed and cataloged by pathologists at McLean Hospital (Belmont, MA, USA). In addition, the severity of pathology in the HD brains was determined using the Vonsattel grading system (Vonsattel et al., 1985).

### Example Dataset 2

This study used a total of 911 individual data from the Alzheimer’s Disease Neuroimaging Initiative (ADNI) (adni.loni.usc.edu). The participants underwent multimodal brain imaging evaluations, including amyloid PET, tau PET and/or structural MRI. The ADNI was launched in 2003 as a public-private partnership, led by Principal Investigator Michael W. Weiner, MD. The primary goal of ADNI has been to test whether serial magnetic resonance imaging (MRI), positron emission tomography (PET), other biological markers, and clinical and neuropsychological assessments can be combined to measure the progression of mild cognitive impairment (MCI) and early Alzheimer’s disease (AD).

In a subset of 744 participants, the Affymetrix Human Genome U219 Array (www.affymetrix.com) was used for gene expression profiling from blood samples. Peripheral blood samples were collected using PAXgene tubes for RNA analysis (Saykin et al., 2015). The quality-controlled GE data includes activity levels for 49,293 transcripts. Molecular PET and MRI images quantifying seven different biological properties were mapped *in vivo* using the following techniques: structural MRI (for structural tissular properties; N=911), Fluorodeoxyglucose PET (for glucose metabolism; N=799), Florbetapir PET (for Aβ deposition; N=906), Arterial Spin Labeling (ASL, for cerebral blood flow; N=341), resting functional MRI (for neuronal activity at rest; N=186), ^18^F-AV-1451 PET (for tau deposition; N=266), and diffusion weighted MRI (for structural brain connectivity; N=128). The preprocessing of the imaging data have been previously described in (Iturria-Medina et al., 2018). For the first six mentioned imaging modalities, representative regional values were calculated for 78 regions covering all the grey matter (Klein and Tourville, 2012). The diffusion weighted MRI data was employed for whole brain region-region structural connectivity (connectome) mapping. All the participants were also characterized cognitively using the mini-mental state examination (MMSE), a composite score of executive function (EF), a composite score of memory integrity (MEM) (Gibbons et al., 2012), and Alzheimer’s Disease Assessment Scale-Cognitive Subscales 11 and 13 (ADAS-11 and ADAS-13, respectively). Also, they were clinically diagnosed at baseline as healthy control (HC), early mild cognitive impairment (EMCI), late mild cognitive impairment (LMCI) or probable Alzheimer’s disease patient (LOAD).

See Table S1 for the corresponding demographic characteristics.

### Procedure

#### Installing *NeuroPM-box* О TIMING 5 - 10 min

##### Δ CRITICAL STEP

To run *NeuroPM-box* on Windows, Linux (or OS X) or Mac systems, you will need either ‘MATLAB 2019b’, or ‘MATLAB Runtime’. Please ensure the Runtime version corresponds to the MATLAB version used by *NeuroPM-box* (i.e. 2019b). MATLAB Runtime can be downloaded for free from: https://www.mathworks.com/help/mps/qs/download-and-install-the-matlab-compiler-runtime-mcr.html.

a. Download the software from neuropm-lab.com/software
b. *For Windows*: run the provided *NeuroPM-box_installer.exe* file to install the software.
c. *For Linux*: call the startup script with the path to MATLAB or the Runtime root folder as an argument:

~~~
>> *MATLAB installed:. /run_NeuroPMbox.sh /*matlabroot*/matlab19b*
>> *Runtime installed:. /run_NeuroPMbox.sh /*mcrroot*/matlab19b_runtime/v94*
~~~ The root folder can be found in MATLAB by checking the variable ‘matlabroot’.
d. *For Mac*: Run the provided *NeuroPM-box_installer.app* file to install the software.

### Executing cTI algorithm О TIMING 5 - 15 min

#### Input data should be

a. a. txt file, with row values representing observations and columns representing features.
b. an ESM file. For each subject, a pseudo-time value (and trajectory position) will be calculated based on her/his baseline data (the unifactorial data; e.g. amyloid, tau). Regional values at baseline will be considered the features on the cTI analysis.
c. a MCM file. Unless specified, all the biological factors (imaging modalities) available will be considered on the cTI (the user can specify if, by the contrary, would like to focus on only one factor/modality).

#### Dimensionality reduction method

in cTI, data exploration and visualization are performed via cPCA (Abid et al., 2018). By controlling the effects of characteristic patterns in the background (e.g. pathology-free and spurious associations, noise), cPCA and its nonlinear version cKernelPCA (Abid et al., 2018) allow visualizing specific data structures missed by popular data exploration and visualization methods (e.g. PCA, Kernel PCA, t-SNE, UMAP). The user can select between “cPCA”, “cKernel PCA”, or “cPCA after applying a smoothing Kernel” to the data (which may reduce the influence of outliers). Of note, before the contrasted dimensionality reduction, all the data features will be ‘boxcox’ transformed (see https://www.ime.usp.br/∼abe/lista/pdfQWaCMboK68.pdf), centered to have mean 0 and scaled to have standard deviation 1.

#### Features preselection

For high-dimensional datasets (e.g. considerably more features than observations), it is necessary to perform an initial selection of features most likely to be involved in a trajectory across the entire population. By default, we apply the unsupervised method proposed by (Welch et al., 2016), which does not require prior knowledge of features involved in the process. Features are scored by comparing sample variance and neighborhood variance. A threshold is applied to select those features with higher score, e.g. you can keep the features with at least a 0.95 probability of being involved in a trajectory (i.e. around 5 % percent of your features dimensionality). Please select the fraction of features that should be used by the cTI algorithm.

#### Background population

The cTI algorithm detects enriched patterns in the population of interest while adjusting by confounding components in the background population (i.e. subjects free of the main effect of interest). To define the background population, the user should provide the list of corresponding IDs, which can be entered by just copying the IDs in the interface, or by loading a “.txt” file in which each row have an ID. Importantly, if the cTI-data is entered as a “.txt” file, we will take as ID the subject’s position on the data (e.g. subject number 10 in the data, at the row 10 of the data matrix, will have ID = 10). If, by the contrary, the cTI-data is coming from ESM/MCM data structures, the Background IDs must correspond with names of the individual folders in the main ESM/MCM folder.

#### Target population (optional)

By default, all the other subjects not defined as background are taken as the target population. However, the user may be interested in to defining the target with a particular subset of subjects (e.g. individuals notably advanced in a disease process). The algorithm will only use then the defined background and target to estimate the model parameters, while the corresponding transformations will be still applied to all the subjects in the data. To define the optional target population, the user should provide the list of corresponding IDs (following same format that for background population), which can be entered by just copying the IDs in the interface, or by loading a “.txt” file in which each row have an ID. Of note, this option is not valid when using the “cKernel PCA” method.

### Δ CRITICAL STEP

Background and target populations have a strong influence on the cTI method (Abid et al., 2018). We recommend defining these taking in to account the biological process of interest. For instance, when studying a given neurological disorder, ensure that the background subjects are free of the studied pathology and with similar demographic characteristics that the target population. The target may be constituted by an heterogenous population, but, if a subset of subjects with highly similar pathological stages/variants is considerably more abundant than subjects at other stages/variants, this subset could statistically dominate (and bias) the contrasted dimensionality reduction technique. In such cases, we recommend pre-defining the target with an equilibrated compendium of disease stages/variants.

#### Adjusting by Covariables (optional)

covariates can be included for linear data adjustment before the feature preselection (if selected) and the trajectories inference analysis. The covariables should be entered as a .txt file, where rows correspond to observations in the main data, the first column to subjects/observations IDs, and the other columns to different covariables. Of note: If a covariable has less than 7 unique values, it will be considered as categorical, and it will be divided in an equivalent number of variables (e.g. for gender information, where one input variable typically has two unique values [female or male], we will replace this variable by two predictor variables, i.e. one for each gender).

#### Additional parameters

The user can define the maximum number of features obtained from the contrasted dimensionality reduction algorithm. Also, the algorithm allows to identify subsets of subjects following potentially different contrasted sub-trajectories. The user can define the maximum number of possible sub-trajectories.

#### cTI Outputs (saved in the input data’s folder)

a. ‘cTI_IDs_pseudotimes_pseudopaths_’ data_name ‘.txt’: file containing the main cTI outputs. First column corresponds to subjects’ IDs. Second column to individual pseudo-time values (a value per subject). From the third to the last column (as many columns as different contrasted sub-trajectories identified), the sub-trajectory or sub-trajectories to which each subject belongs to. As mentioned, each subject can belong to more than one sub-trajectory (e.g. when two disease variants overlap at their beginning).
b. ‘cTI_cPCs_’ data_name ‘.txt’: obtained contrasted principal components.
c. ‘cTI_weights_’ data_name ‘.txt’: loadings/weights (one column per initial feature).
d. ‘cTI_features_preselected_’ data_name ‘.txt’: when features preselection is performed, this file will contain the indices of those features most likely to be involved in a trajectory across the entire population, and subsequently used in the cTI analysis.

### Executing ESM О TIMING 5 - 25 min per subject

#### Organizing your data for ESM

The software provides an automatic tool to import all the needed data for ESM evaluation (on the main interface, click “Complementary_tools”). The data should be organized individually, with a folder per subject. For compatibility across different models in the toolbox, the images can be organized in the same way that for the multi-modal models (e.g. MCM). In case the software detects multiple imaging modalities, the user will be asked about which one should be used for ESM. Each subject’s folder should include (see Fig. 6):

**Figure 5.**
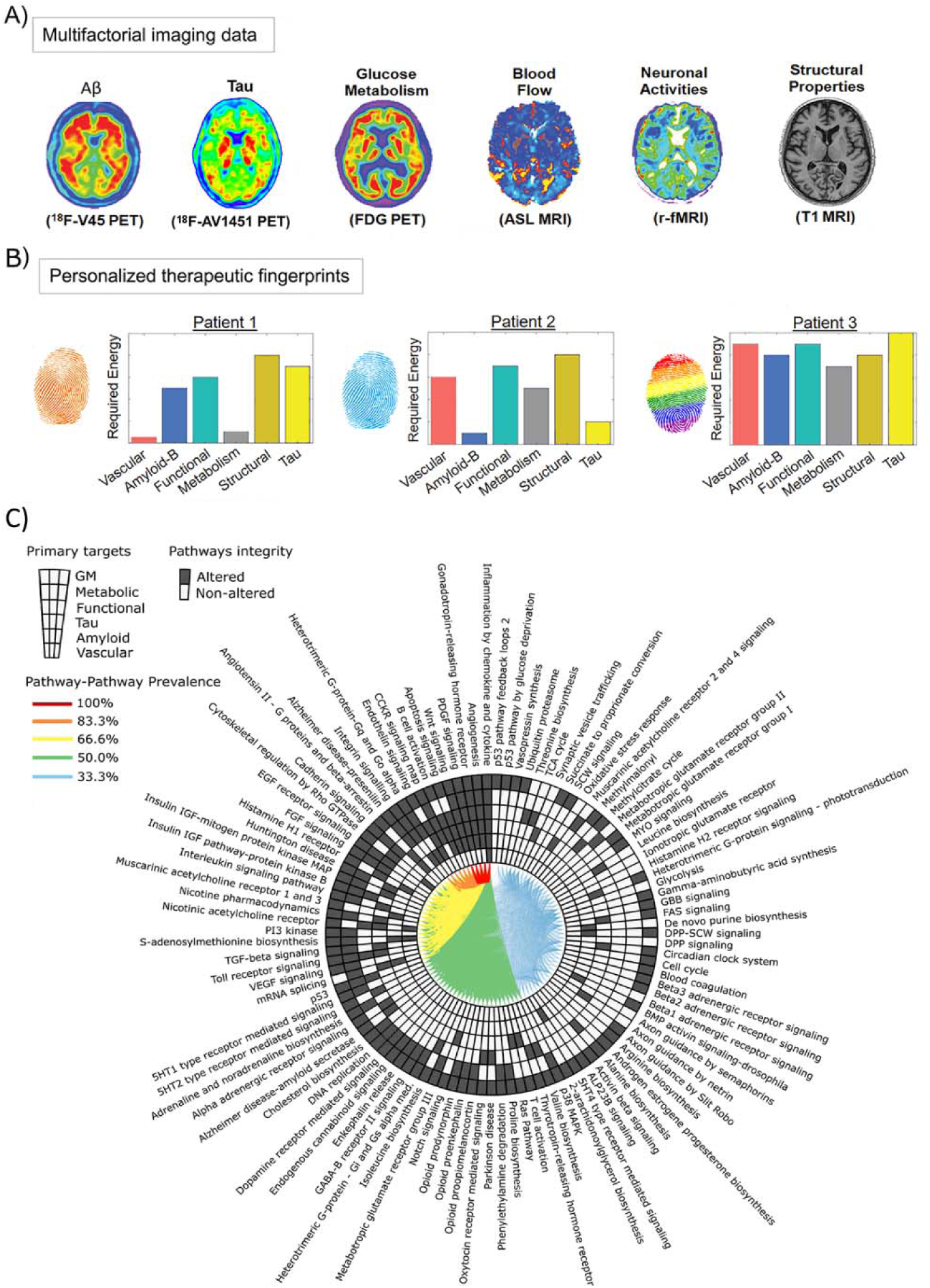
From multimodal imaging to therapeutic fingerprints and altered molecular pathways (ADNI data). Imaging for amyloid, tau, CBF, functional activity at rest, glucose metabolism, and gray matter density. A network-based approach (Iturria-Medina et al., 2017) allows individual characterization of the intra-brain direct factor-to-factor biological interactions and the multifactorial spreading mechanisms through vascular/anatomical connections. Inverting the model’s fundamental equation allows estimation of the changes required to produce a desired state (e.g. healthy). Thus, the pTIF is defined as the set of changes required for each patient. B) Dissimilar pTIF patterns for three participants with the same diagnosis. Note that Patient 1 requires lower cost-energy values for vascular and metabolic interventions, while Patient 2 requires lower values for anti-Aß and anti-tau interventions, suggesting the identification of specific single-target therapies that may benefit these patients (e.g. physical exercise and aducanumab, respectively). However, Patient 3 would requires high cost-energy for all the single-target interventions, suggesting that combinatorial (and not single-target) treatments will be most beneficial in this case. C) Altered molecular pathways (blood data) underlying distinctive single-target therapeutic needs. The pathways were sorted according to their prevalence levels across all the single-target specific subgroups, starting at 12 o’clock and following a counter-clockwise direction. Each link between a given pair of pathways corresponds to the percentage of subgroups for which both molecular pathways were found to be affected. Images adapted with permission from (Iturria-Medina et al., 2018).

**Figure 6.**
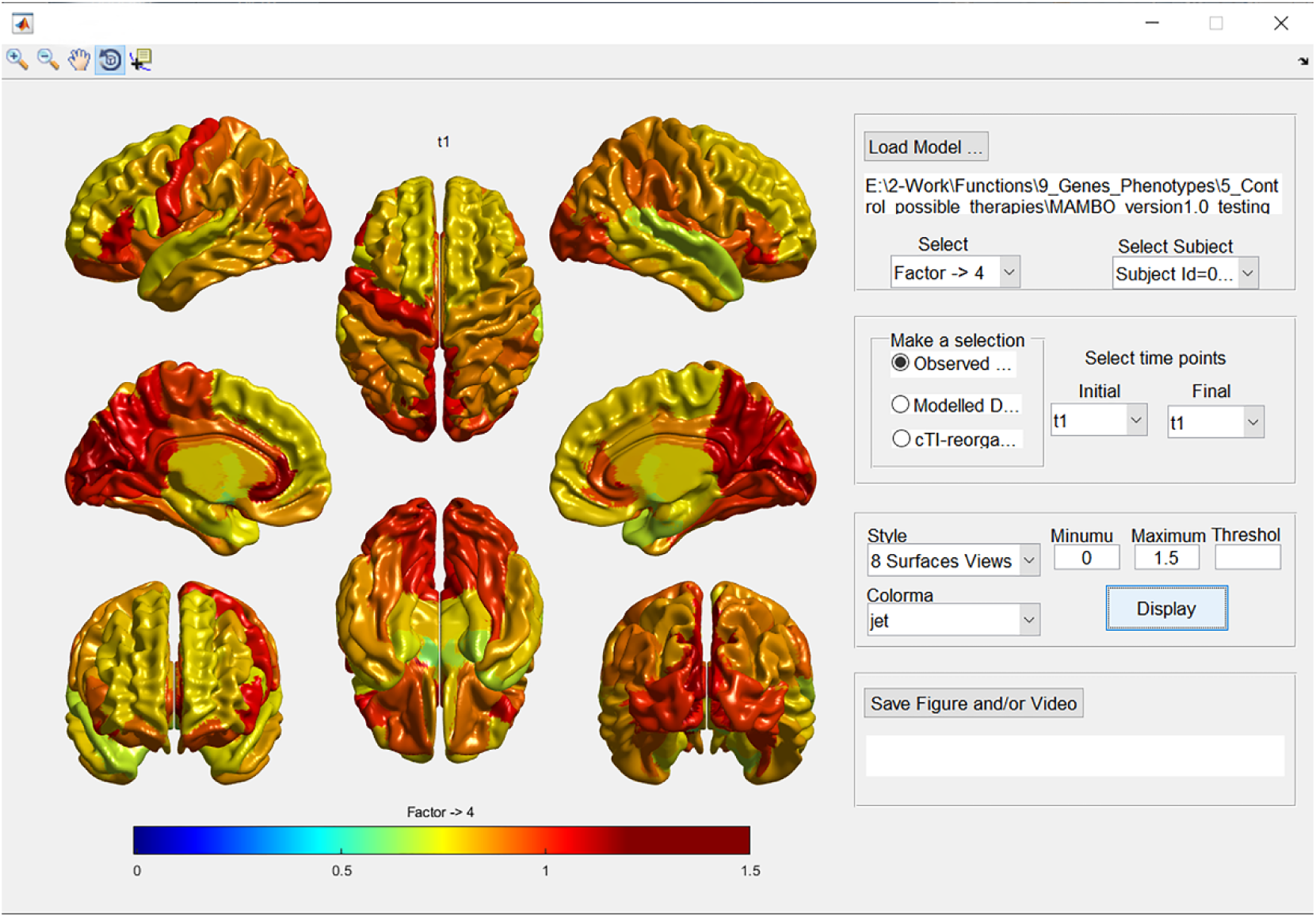
*NeuroPM-viewer* interface depicting glucose metabolism (FDG PET) in an 8 brain surfaces view. In addition to select multiple visualization settings (data-type, time windows, colormaps, minimum and maximum values, threshold), the user can save the depicted figures in traditional images formats (.jpg, .png, and .tif) or dynamic brain videos (.avi).

a. Brain images (.nii or .mnc) corresponding to each biological factor of interest, e.g:

~~~
*factor_1_t0.nii, factor_1_t1.nii, factor_1_t2.nii, factor_1_t3.nii*
*factor_2_t0.nii, factor_2_t1.nii, factor_2_t2.nii,*
*factor_3_t0.nii, factor_3_t1.nii, factor_3_t2.nii, factor_3_t3.nii…*
~~~ Include as many factors and time points as available. Of note, t1, t2, t3… should be numeric values.
b. Gray matter parcellations images (.nii or .mnc), e.g.:

~~~
*GM_parcellation_t0.nii, GM_parcellation_t1.nii, GM_parcellation_t2.nii, GM_parcellation_t3.nii…*
~~~ Importantly, if there is only a common population parcellation at the group level (e.g. coming from other study/template), the parcellation files can be, alternatively, saved at the root folder containing all the subject’s folders. In any case, please be sure to include at least one parcellation image for each subject or for the whole population.
c. Connectomes (mandatory, e.g. anatomical and/or vascular networks) files: Multiple (.txt or .csv) files (one for each available time point, with rows and columns corresponding to brain regions), for example:

~~~
*‘connectome1_t0.txt’, ‘connectome1_t1.txt’, ‘connectome1_t2.txt’…*
~~~ Importantly, if the connectivity information is only available at the group level (e.g. coming from other study/template), the connectome files can be, alternatively, saved at the root folder containing all the subjects’ folders. In any case, please be sure to include at least one connectome matrix, with same number of rows and columns (i.e. regions) that your parcellation.

#### Data standardization (optional)

The voxel or regional values can be standardized at the individual level using user-defined reference regions (e.g. cerebellum). When using this option, the user should define:

a. labels of reference regions, corresponding with the regions’ numeric values in the provided parcellation images.
b. if the reference regions should NOT be removed for the posterior modeling analysis (by default, the references are removed).

#### Probabilistic data standardization (optional)

the original ESM version was defined in probabilistic terms. Although the current implementation can work with raw data values (default), the user can opt to convert the raw voxel values to probabilities, by comparing each voxel with the distribution of maximum in the reference regions.

#### *Reversing scale* (optional)

by default, it is considered that higher signal values in the images will be more reflective of the process of interest (e.g. misfolded proteins deposition quantified with PET imaging). However, for some data modalities (e.g. gray matter density), a lower value would imply a stronger effect (e.g. structural atrophy). For those cases, the user can opt to reverse the data’s scale via a linear scaling (the numerical scale will runs in the opposite direction).

#### Input file for ESM optimization

After using “Organizing input for ESM” in “Complementary Tools”, input the file named “Input_data_…_ESM.mat”, which should be saved in the folder “ESM_data_results” inside your data’s initial directory. All optimization results will be also saved in this file as well as in text files.

#### The Epidemic Spreading Model (ESM) can be optimized at

a. the individual level (only if longitudinal data is available). A minimum of 3 time points is required (subjects without enough data will not be analyzed). Individual parameters (clearance, production, onset time) will be saved as a .txt file where rows are subjects and columns are IDs, effective clearance, effective production and onset time (if required). Additionally, for visualization purposes, these variables will be saved in the original “_ESM.mat” file.
b. the population level. **Δ CRITICAL STEP** First, use the “contrastive Trajectories Inference (cTI)” algorithm, where a pseudo-time value will be obtained for each subject based on his/her baseline data (see instructions about). Alternatively, use your own subjects’ stratification, coming from using cTI on a different data modality (e.g. molecular, clinical) or from a different computational method (for using this option, see “Adding new stratification to ESM/MCM files” in “Complementary Tools”). Then, the subjects will be ordered according to their characteristic pseudo-path and pseudo-time, constituting a pseudo-longitudinal data for ESM optimization. Sub-group parameters (clearance, production, onset time) will be saved in a .txt file, where rows will be sub-groups (each corresponding to subjects belonging to a characteristic pseudo-path), and columns will have: subject(s) ID, effective clearance, effective production and onset time (if required). Additionally, for the visualization purposes, these variables will be saved in the original “_ESM.mat” file.

#### Production and clearance factors can be defined as

a. sigmoidal (default): following sigmoid functions that depend on global production, or clearance rates, and on the current regional value. In this case, each brain region has its own effective production or clearance rate, which may change with time. The effective regional production is assumed to increase with the local signal (e.g. the more amyloid a region has, the higher chance that it will produce and spread more amyloid seeds). Contrary, effective clearance is assumed to decrease with the local signal (i.e. more “infected” regions are less able to clean the accumulated “agents”).
b. constants, with same rate across all the spreading process.

#### Epicenters identification

The spreading process under study is assumed to start in a set of specific brain regions, from which the “agent” propagates across physical brain connections. If the epicenter regions are known, they can be predefined by the user and the optimization algorithm will continue from these. Otherwise, the epicenter(s) will be estimated as the region[s] with the highest value[s] at the onset time. Both backward and forward integration procedures will be used for estimating the most likely epicenter(s), using the maximum number of allowed epicenter regions defined by the user.

### Δ CRITICAL STEP

When working with some imaging modalities (e.g. SUVr PET), a nonzero regional value doesn’t necessarily implies the presence of the studied “agent” (e.g. amyloid or tau deposition) but just background fluctuations on the image signal (determined, for example, by different physiological factors and/or random noise). Commonly, in literature, a positivity threshold is applied to detect “agent” presence or not. Consequently, for estimating the regional epicenters, our algorithm allows to predefine a maximum (nonzero) value below which the regions are still considered free of “agent” presence but with their typical background ‘noise’. Only regions over this value will be considered as likely epicenters (and, correspondingly, all regions below are considered non-epicenters). The user may see some improvement in model fit when using a nonzero value, due to the non-epicenters will not be forced to be zero at the onset-time (i.e., increasing correspondence with reality, in which regions may never have an exact zero value, depending on the imaging modality used).

Of note, increasing ‘too much’ the maximum value for non-epicenters may result in data overfitting, because the non-epicenters may have a large enough range, and the corresponding numerical flexibility, to adapt to the studied process. That is, if the “agent” positivity threshold is not conservative enough (i.e. too high), all brain regions can potentially behave as epicenters. We suggest a value about or below the 5% of the typical maximum value in the analyzed imaging modality (or about/below 0.05 if working with probabilistic values).

#### Estimating Onset time

this measure should be interpreted as the time at which the intra-brain spreading process under study started (e.g. the age at which amyloid and tau proteins appeared and started propagating in the brain). If known, the user can provide the onset time. Otherwise, it will be estimated by the optimization algorithm. In such a case, the user would need to provide the minimum possible value (default: zero).

#### ESM Outputs

(saved as .txt files in the folder “ESM_data_results” inside your data’s initial directory, and as MATLAB variables in the ESM’s .mat file):

a. ‘ESM_subject_…_ACCURACY.text’: model accuracy (in %).
b. ‘ESM_subject_…_resnorm.text’: 2-norm of the residuals.
c. ‘ESM_subject_…_parameters.text’: obtained model parameters (production and clearance, respectively) in their original numeric scale.
d. ‘ESM_subject_…_effective_production.text’: production and clearance model parameters may be difficult to interpret in the original numeric scale. To facilitate parameter comparability across subjects, the individual production parameter’s marginalized across all possible “agent” concentration/probability values, obtaining the effective individual production rate.
e. ‘ESM_subject_…_effective_clearance.text’: similarly, the individual clearance parameter’s marginalized across all possible regional concentration/probability values, obtaining the effective individual clearance rate.
f. ‘ESM_subject_…_sorted_most_likely_epicenters.text’: when the epicenter regions are not provided by the user, a list of most likely epicenters will be provided, based on the model’s optimization.
g. ‘ESM_subject_…_S0.text’: obtained “agent” concentration/probability values at the estimated onset time, i.e. initial perturbation triggering the spreading process. Notice that the solution is not necessarily sparse (i.e. all regions could have a non-zero value), but those regions with highest values (over a positivity threshold) should be considered the most likely propagation epicenters.
h. ‘ESM_subject_…_onset_time.text’: estimated (or provided) time at which start the intra-brain “agent” spreading process.
i. ‘ESM_subject_…_simulated_data.text’: by default, the optimized model parameters will be used to generate/simulate 30 data points, equally positioned in time from: the estimated (or provided) onset time, until the last available time point plus the half of the longitudinal time window of the subject’s real data. In addition, the model will generate the data at the observed time points, simulating a total of N_times_ = 30 + subject time points. This simulated data (saved here as a [N_regions_*N_modalities_ x N_times_] matrix) can be of particular interest to visualize spatiotemporal brain changes in a continues time scale. Also, the generated data at the observed time points can be used for model validation, comparing with the real observed data.
j. ‘ESM_subject_…_simulated_times.text’: corresponding time values to the generated/simulated data.

For the case when the model is applied at the population level (after using the pseudo-times and sub-trajectories from *cTI*), all the same model outputs will be saved for each previously identified sub-trajectory/subgroup, e.g. ‘ESM_Subgroup_1_ACCURACY.text’ and ‘ESM_Subgroup_2_ACCURACY.text’.

### Executing MCM О TIMING 5 - 25 min per subject

#### Organizing your data for MCM

The software provides an automatic tool to import all the needed data for MCM evaluation (on the main interface, click “Complementary_tools”). The data should be organized individually, with a folder per subject, each subject’s folder including (see Fig. 8):

**Figure 7.**
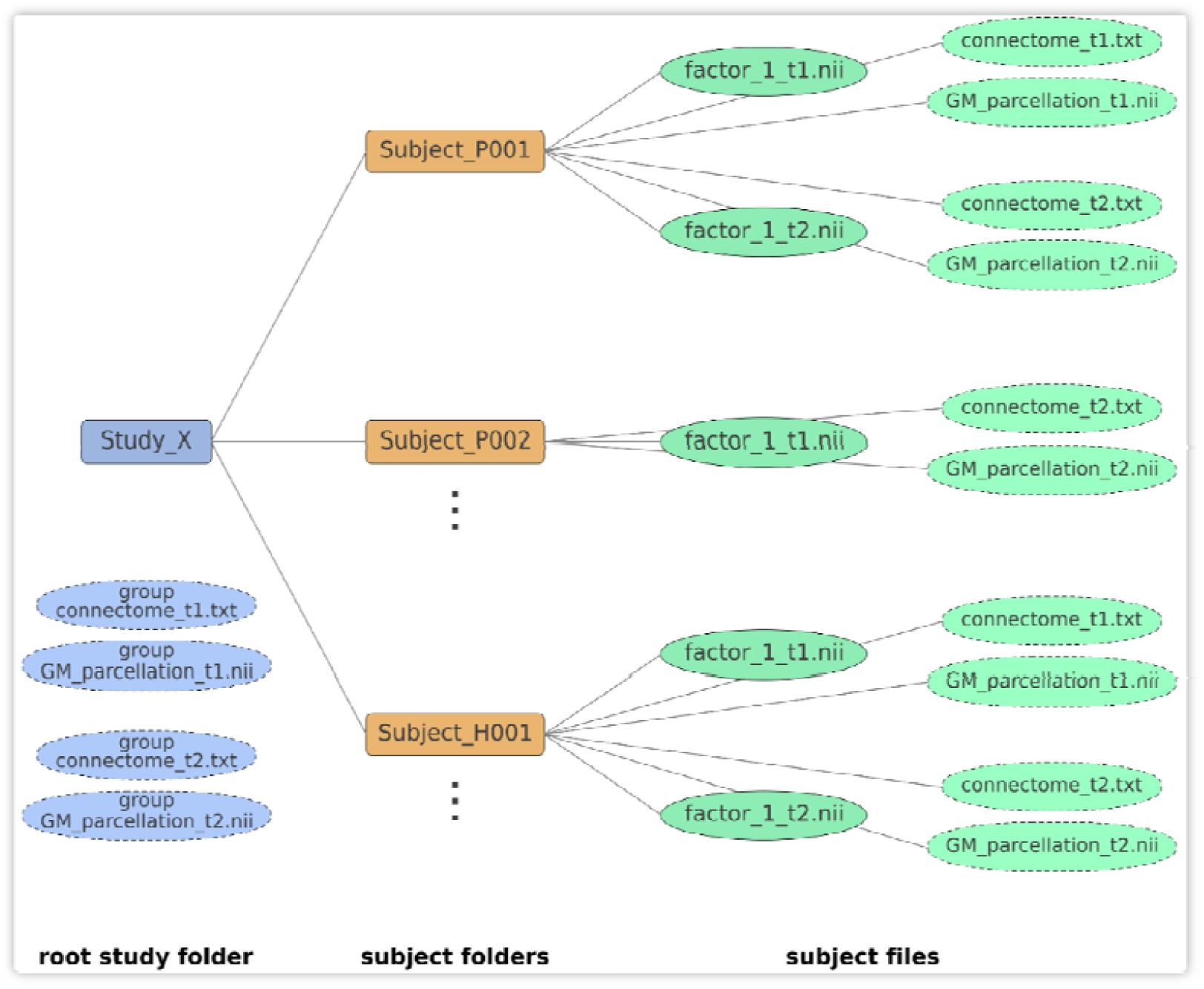
Schematic for data organization in an ESM study. As mentioned, if the brain parcellation(s) and/or connectivity information are only available at the group level (e.g. coming from a template or another study), the parcellation(s) and/or connectome files can be, alternatively, saved at the root folder containing all the subjects. Please be sure to include at least one connectome matrix for the first modality, with same number of rows and columns (i.e. regions) that your parcellation.

**Figure 8.**
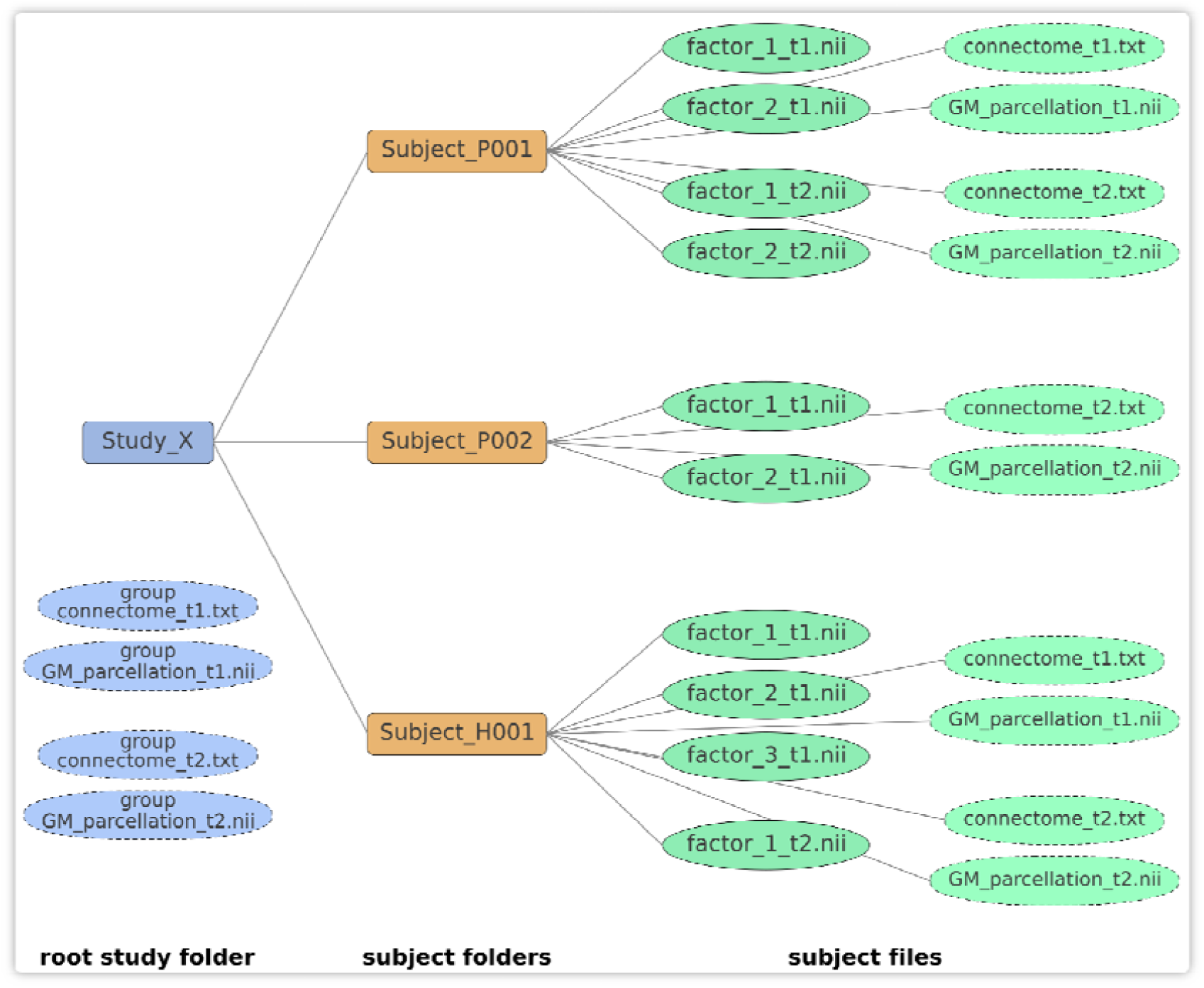
Schematic for data organization in an MCM study. As mentioned above, if the brain parcellation(s) and/or connectivity information are only available at the group level (e.g. coming from a template or another study), the parcellation(s) and/or connectome files can be, alternatively, saved at the root folder containing all the subjects. Please be sure to include at least one connectome matrix for the first modality, with same number of rows and columns (i.e. regions) that your parcellation.

a. Brain images (.nii or .mnc) corresponding to each biological factor of interest, e.g.:

~~~
*factor_1_t0.nii, factor_1_t1.nii, factor_1_t2.nii, factor_1_t3.nii factor_2_t0.nii, factor_2_t1.nii, factor_2_t2.nii*,
*factor_3_t0.nii, factor_3_t1.nii, factor_3_t2.nii, factor_3_t3.nii*
~~~ Include as many factors and time points as available. t1, t2, t3… should be numeric values.
b. Gray matter parcellations images (.nii or .mnc):

~~~
*GM_parcellation_t0.nii, GM_parcellation_t1.nii, GM_parcellation_t2.nii, GM_parcellation_t3.nii…*
~~~ Importantly, if there is only a common parcellation at the group level (e.g. coming from another study/template), the parcellation files can be, alternatively, saved in the root folder containing all the subjects’ folders. In any case, please be sure to include at least one parcellation image for each subject or for the whole population.
c. Connectomes (mandatory, e.g. anatomical and/or vascular networks) files Multiple (.txt or .csv) files (one for each connectome modality and each time point, with rows and columns corresponding to brain regions), named for example:

~~~
*connectome1_t0.txt, connectome1_t1.txt, connectome1_t2.txt…*
~~~ Optionally, if a second connectome modality is available, name it as:

~~~
*connectome2_t0.txt, ‘connectome2_t1.txt, connectome2_t2.txt…*
~~~ Importantly, if the connectivity information is only available at the group level (e.g. coming from other study/template), the connectome files can be, alternatively, saved at the root folder containing all the subjects’ folders. In any case, please be sure to include at least one connectome matrix for the first modality, with same number of rows and columns (i.e. regions) that your parcellation.
d. Output variables file (optional, e.g. cognitive/clinical evaluations): In each subject’s folder, include a file named ‘outputs_variables.txt’, with as many rows as time points, and organized as

~~~
*value_variable1 value_variable2* (*…*) *value_variableN evaluation_t0*
*value_variable1 variable2* (*…*) *value_variableN evaluation_t1*
~~~ Include as many time points as available. For each missing value, use a ‘NaN’.
e. External Inputs file (optional, e.g. drugs intake): In each subject’s folder, include a file named ‘input_variable.txt’, with as many rows as time points or inputs presented, and organized as

~~~
*starting_time1 finishing_time1 value_input_intensity* (*constant value if not changing*) *specific_target_regions* (*numbers in GM parcellation, leave empty if all regions*)
*starting_time2 finishing_time2 value_input_intensity specific_target_regions*
*starting_time3 finishing_time3 value_input_intensity specific_target_regions*
~~~ Include as many time points as available. All entrances should be numeric values. *value_input_intensity* should be a constant value reflecting the intensity of the stimulus in the specified time window. specific_target_regions should be the numbers of the targeted regions in the GM parcellation, leave empty if all regions were targeted.

### Δ CRITICAL STEP

When working with longitudinal imaging data, it is common to have subjects with missing time points and/or imaging modalities. During the data’s organization, the user can opt to impute the missing data via the trimmed scores regression algorithm (Folch-Fortuny et al., 2016). We strongly recommend using this option, particularly when the subjects may have different acquired time points and imaging modalities (e.g. ADNI data). Otherwise, be sure to have a complete dataset for each subject. If imputation is not selected, subjects with missing imaging modalities will be removed from the analysis.

#### Input file for MCM

an MCM file. After using “Organizing input for MCM” in “Complementary Tools”, you should have a file for MCM evaluation/optimization, termed “Input_data_…_MCM.mat”, and saved in the folder “MCM_data_results” inside your initial data’s directory. All optimization results will be also saved in this file as well as in text files.

#### The MCM can be optimized at

f) the individual level (only if longitudinal data is available). A minimum of 3 time points is required (subjects without enough data will not be analyzed). Individual parameters (e.g. factor-to-factor causal interactions) will be saved as a .txt file where rows are subjects and columns are subject ID and parameters. Additionally, for visualization purposes, these variables will be saved in the original “_MCM.mat” file.
g) the population level. **Δ CRITICAL STEP** First, use the “contrastive Trajectories Inference (cTI)” algorithm, where a pseudo-time value will be obtained for each subject based on his/her baseline data (see Iturria-Medina et al., 2019). Alternatively, enter your own subjects’ stratification, coming from using cTI on a different data modality (e.g. molecular, clinical) or from a different computational method (for using this option, see “Adding new stratification to ESM/MCM files” in “Complementary Tools”). Then, the subjects will be ordered according to their characteristic pseudo-path and pseudo-time, constituting a pseudo-longitudinal data for MCM optimization. Sub-group parameters (e.g. factor-to-factor causal interactions) will be saved in a .txt file where rows are sub-groups (each corresponding to subjects belonging to a characteristic pseudo-path) and columns are subject ID, and model parameters. Additionally, for visualization purposes, these variables will be saved in the original “_MCM.mat” file. Please see also the “contrastive Trajectories Inference (cTI)” interface.

#### Estimating initial system perturbation before baseline

Use this option if you consider that the subjects may have an underlying “alteration” process started before their first imaging evaluation (e.g. a long-term neurodegenerative process started years time before the baseline). Importantly, do you know the clinical state of the subjects before the first “alteration”? Does any of the subjects in the current population is “free” of relevant dynamic changes (at least until their first evaluation)? If so, those subjects can be considered “controls” and, by specifying their IDs in a .txt file, you can help the model to have an approximation of the data’s typical distribution before the potential perturbation under study occurred. If this information is available, we strongly recommend loading their IDs (i.e. the corresponding folder names of those subjects). Otherwise, the first evaluation will be taken as reference for quantifying potential brain alterations/perturbations, for each subject.

#### Estimating initial system perturbation after baseline

If you have information on the time when an external event/input perturbed the brain system, we recommend including this information in the data’s organization. For MCM optimization, use then the “Known event(s) or input(s)” option, which will consider the corresponding information. If not, select “Unknown event(s) or input(s)”. If “Estimate S0” is also selected in the “Optimization” panel, the algorithm will estimate the initial system perturbation.

### Δ CRITICAL STEP

By default, we use a spline-based optimization method to solve the system of differential equations (Ramsay, 2006; Ramsay et al., 2007). This results in a very fast optimization, without needing to propose initial parameters. Alternatively, if this option is not active, we will use a conventional gradient-based optimization method (trust-region-reflective algorithm, (Coleman and Li, 1996, 1992)), which is started at multiple seed points for avoiding local minimum solutions. The former method takes from seconds to a few minutes per subject, while the latter method results in a significantly larger computational time, in the order of hours per subject. In our data, both methods provided comparable results.

#### Considering external inputs (optional)

Select this option to estimate the effects of any known external input on the brain system. For this, you should have previously included information about the external input in the data’s organization process (see details above). The optimization algorithm will estimate a global measure of the impact that the input has on each considered biological factor.

#### Estimate S0 (optional)

Use this option if you would like to obtain an estimate of the initial perturbation on the brain system (i.e. what may have caused the initial propagation of biological alterations on the system).

#### Regularization (optional)

If you are using the “trust-region-reflective” algorithm (and not the spline smoothing), a Tikhonov regularization will be used during the parameters’ optimization. The regularization may significantly improve the parameters, its robustness and biological interpretability, although it will also require a considerably larger computational time.

#### Parallel calculus (optional)

When using the “trust-region-reflective” algorithm (and not the spline smoothing), use this option if you would like to use your PC’s multiple cores during model optimization. It may result in a significant reduction of the computational time.

#### Number of iterations (optional)

In order to refine the output parameters, the MCM is optimized multiple times. The higher the number of iterations, the longer the computational time, but potentially better results.

#### MCM Outputs

(saved as .txt files in the folder “MCM_data_results” inside your data’s initial directory, and as MATLAB variables in the MCM’s .mat file):

Of note, here we will refer to N_regions_ and N_modalities_ as the number of brain regions and imaging modalities considered, respectively. Each biological factor corresponds to a given imaging modality.

a. ‘MCM_subject_’ subject_ID ‘_accuracy_resnorm.txt’: obtained model accuracy (in %) and 2-norm of the residuals, respectively.
b. ‘MCM_subject_’ subject_ID ‘_parameters.txt’: obtained model parameters in their original numeric scale. All parameters are also saved as different outputs according their specific role and biological interpretation on the model (see descriptions below). First N_modalities_*N_modalities_ corresponds to direct factor-to-factor interactions (rows are *seeds*, columns are *targets*; see *Effective causality* below). Second N_modalities_ elements correspond to factor-specific scaling/weighting values associated to the intra-brain spreading processes (a high value for factor *m* suggesting an strong role of the intra-brain spreading process for this factor; however, for *post hoc* analysis, we recommend to use instead the *Effective spreading* output described below). Next N_modalities_ elements reflect the fraction [0,1] of factor/modality specific alterations spreading through the first brain network specified (e.g. if the user provided both anatomical and vascular brain connectomes, these output parameters would be reflecting the factor-specific fraction of spreading via the anatomical network, while the difference with 1 would reflect the fraction of spreading by the vascular network). Finally, if external input information’s specified, the last N_modalities_ parameters will correspond to the global direct influence of the input signal on each factor/modality considered (see also ‘intervention_effects’ and ‘relative_intervention_effects’ outputs, described below).
c. ‘MCM_subject_’ subject_ID ‘_Effective_causality.txt’: relative direct factor-to-factor influences (effective causal effects). Square matrix of size [N_modalities_ x N_modalities_], where the element *n,m* corresponds to the relative direct effect of factor *n* over *m* (n→m) while accounting for all other factors interactions and intra-brain spreading, that is, the percent of regional changes in factor *m* that are caused by the direct influence of factor *n*. It is calculated as 100 multiplied by the sum of the direct effects of *n* over *m*, across all brain regions, relative to the sum of direct effects of all the biological factors over *m* (including itself) and spreading effects.
d. ‘MCM_subject_’ subject_ID ‘_Effective_X_initial.txt’: initial estimation of the relative factor-to-factor influences (effective causal effects), obtained before the model’s optimization via a regression analysis and only used as *a priori* input for model estimation (not recommended to be used in *post hoc* analysis).
e. ‘MCM_subject_’ subject_ID ‘_Effective_spreading.txt’: relative spreading of considered factors, where the element *m* reflects the percent of the spatiotemporal changes in factor *m* that are presumably caused by its spreading through brain physical connections (and not by factor-to-factor interactions or external inputs).
f. ‘MCM_subject_’ subject_ID ‘_Effective_incoming.txt’: relative incoming influences for considered factors. Similar to the in-strength measure in a directed network, element *m* reflects the percent of regional changes in factor *m* that are caused by the direct influences of all the other biological factors, excluding self-effects. This measure allows the identification of the most vulnerable and influenced biological factors (reflected in imaging modalities) during a given brain process.
g. ‘MCM_subject_’ subject_ID ‘_Effective_outgoing.txt’: relative outgoing influences for considered factors. Similar to the out-strength measure in a directed network, reflects the percent of regional changes in all the considered biological factors that are caused by the direct influence of a given biological factor *n*, excluding self-effects. This measure can be particularly useful to detect the most influential biological factors during a brain process.
h. ‘MCM_subject_’ subject_ID ‘_A_networks_optimum.txt’: identified *multifactorial causal network* (matrix *A* in Iturria-Medina et al., 2017, *Neuroimage*), with parameters controlling regional multifactorial causal interactions and effects propagation through physical networks (e.g. axonal and vascular connectomes). Matrix of size [N_regions_*N_modalities_ X Nregions*Nmodalities].
i. ‘MCM_subject_’ subject_ID ‘_initial_perturbation.txt’: estimated (or provided by the user if external input information’s specified) initial system perturbation, i.e. a vector of size [N_regions_ x N_modalities_], where first N_regions_ elements correspond to perturbations in imaging modality/factor 1, second N_regions_ elements to perturbations in modality/factor 1, and so on, until.
j. ‘MCM_subject_’ subject_ID ‘_perturbation_time.txt’: estimated (or provided by the user if external input information’s specified) time at which happens the initial system perturbation.
k. ‘MCM_subject_’ subject_ID ‘_intervention_effects.txt’: if external input information’s specified, this output consist of N_modalities_ parameters corresponding to the estimated global direct influence of the input signal on each factor/modality considered (see also ‘relative_intervention_effects’ below).
l. ‘MCM_subject_’ subject_ID ‘_relative_intervention_effects.txt’: same that ‘intervention_effects’, but after normalizing each factor *m*’s corresponding value by the sum of direct effects of all the biological factors over *m* (including itself), spreading- and input-effects.
m. ‘MCM_subject_’ subject_ID ‘_est_data_before_perturbation.txt’: vector with N_regions_*N_modalities_ elements corresponding to estimated multifactorial regional values at the time that the initial perturbation occurred.
n. ‘MCM_subject_’ subject_ID ‘_simulated_data.txt’: by default, the optimized model parameters will be used to generate/simulate 30 multifactorial data points, equally positioned in time from: the estimated (or provided) onset time, until the last available time point plus the half of the longitudinal time window of the subject’s real data. In addition, the model will generate the data at the observed time points, simulating a total of N_times_ = 30 + subject time points. This simulated data (saved here as a [N_regions_*N_modalities_ x N_times_] matrix) can be of particular interest to visualize spatiotemporal brain changes in a continues time scale. Also, the generated data at the observed time points can be used for model validation, comparing with the real observed data.
o. ‘MCM_subject_’ subject_ID ‘_simulated_times.txt’: corresponding time values to the generated/simulated data.

For the case when the model is applied at the population level (after using the pseudo-times and sub-trajectories from *cTI*), all the same model outputs will be saved for each previously identified sub-trajectory/subgroup, e.g. ‘MCM_Subgroup_1_ACCURACY.text’ and MCM_Subgroup_2_ACCURACY.text’.

### Executing pTIF О TIMING 10 - 30 min per subject

#### Input data for pTIF estimation

the MCM’s .mat file after optimization. See “Organizing input for MCM” in “Complementary Tools”, and *Executing MCM* subsection above.

#### The pTIF can be estimated when

a. having individual longitudinal data. Firstly, the MCM approach should be optimized at the individual level, using the available longitudinal data. Then, each individual multifactorial causal network will be analyzed, depending on the selected options (see below) to provide an individual pTIF (a vector with the required energy deformations to move a subject’s state at the time of her/his final evaluation to a desired state). The corresponding pTIF values (and the chosen options) will be saved as “…pTIF.txt” in the MCM data/results folder.
b. having cross-sectional data for a relatively large population. **Δ CRITICAL STEP** First, use the “contrastive Trajectories Inference (cTI)” algorithm with your MCM file. Then, the subjects will be ordered according to their characteristic pseudo-path and pseudo-time, constituting a pseudo-longitudinal data for MCM optimization. Once the (sub)population MCM is optimized, the group’s multifactorial causal network will be analyzed, depending on the selected options (see below), to provide an individual pTIF (a vector with the required energy deformations to conduce a subject’s state at the time of the evaluation to a desired state). The temporal analysis will be based on the pseudo-time scale. All the individual pTIF values (and the chosen options) will be saved as “…pTIF.txt” in the MCM data/results folder.

#### Control brain factors or output variables

The MCM can be used as *in silico* evaluator of external inputs, which can focus on obtaining a desired state following two different control strategies:

a. *Full control*: focuses on controlling all considered brain factors and regions.
b. *Output control*: focuses on controlling cognitive/behavioral states, without necessarily modifying all the studied brain properties, and focusing on a specific output variable (e.g. a given cognitive metric).

#### Desired system state

When estimating the optimum signal to conduce the brain system or its outputs from a current state (last time point) to a desired state, the user should specify the final desired state, opting to:

a. keeping stable the observed alterations (desired state is equal to initial state).
b. reducing the current state’s alterations to a given percent (provide a value between 0 [reducing all alterations to zero level] and 100 [causing no change]).
c. reducing the current state’s alterations to the mean level of the control subjects, which is the reference for calculating the alterations (this analysis can only be performed if control subjects were defined before MCM optimization).

#### Intervention duration

Time window of the hypothetical brain intervention. **Δ CRITICAL STEP** It must be in the same scale that the reported time in the observed data. However, if using a pseudo-time metric instead, be sure to always enter a value between 0 and 1. In this case, it is important to consider that the interval 0 to 1 may be equivalent to the whole time that takes the studied process (e.g. a disease covering years of progression). We recommend using a conservative number, for example, if a disease under study usually takes 10 years to develop, and you would like to simulate a 1-year intervention, define an intervention duration equal to 1/10.

#### Targeting combinations of factors

Define the number of factors (imaging modalities) that are going to be targeted during the hypothetical external intervention. For combinatorial interventions, we recommend combinations of as many biological factors as available imaging modalities.

#### pTIF Outputs

(saved as .txt in the input data’s folder and as variables in the MCM’s .mat file). Here we will refer to N_subjects_, N_regions_ and N_modalities_ as the number of subjects, brain regions and imaging modalities considered, respectively. Each biological factor corresponds to a given imaging modality.

a. ‘Results_Input_data_(…)_MCM_OPTIONS_for_pTIF.txt’: options selected by the user for pTIF estimation (e.g. control strategy, input duration, combinations of factors).
b. ‘Results_Input_data_(…)_MCM_pTIF.txt’: global pTIF matrix of size [N_subjects_ x N_pTIF_]. The individual pTIF corresponds to a numeric multivariate vector with dimensionality determined by all possible “tested” interventions. The number of pTIF elements (N_pTIF_) depend on the number of imaging modalities/factors considered and the number of factors combined (see *Targeting combinations of factors* option above). For instance, for N_modalities_=6, the number of all possible single-target or combinatorial interventions (up to a maximum of 6 factors) is 63, which finally defines the individual pTIF vector.
c. ‘Results_Input_data_(…)_MCM_regional_pTIF.txt’: regional pTIF matrix of size [N_subjects_ x N_regions_*N_pTIF_]. For each “tested” intervention, the global pTIF value (reported in the described global pTIF matrix, above) corresponds to the sum of all the energy-cost regional values required to cause the desired change. Contrary, the regional pTIF matrix include all the regional values before adding them, which provides a detailed characterization of the estimated factor(s)-specific changes for each brain region.

## Data Availability

All data used in this study are publicly available at the Gene Expression Omnibus (GEO; ncbi.nlm.nih.gov/geo/, accession number GSE44772) and the Alzheimer's Disease Neuroimaging Initiative (ADNI; adni.loni.usc.edu).

https://www.neuropm-lab.com/neuropm-box.html

## Software availability

User-friendly *NeuroPM-box* standalone applications for Windows, Linux and Mac environments are freely available for academics and non-for-profit researchers at neuropm-lab.com/software. A detailed PDF software tutorial is also provided.

## Acknowledgments

We want to give a special thanks to Ms. Sheila Linden for her generous funding support to this project. We also thank Joanne Clark (Executive Director of the *Ludmer Centre for NeuroInformatics and Mental Health* at McGill), for her support and valuable comments about the software, tutorial and manuscript. This project was also undertaken thanks in part to the following funding awards to YIM: the *Fonds de la Recherche en Sante du Quebec* (FRQS) Research Scholars Junior 1, the Weston Brain Institute Rapid Response AD program 2018, and the New Investigator start-up grant from McGill University’s *Healthy Brains for Healthy Lives Initiative* (*Canada First Research Excellence Fund*). In addition, dataset-2 collection and sharing for this project was funded by ADNI (National Institutes of Health Grant U01 AG024904) and DOD ADNI (Department of Defense award number W81XWH-12-2-0012). ADNI is funded by the National Institute on Aging, the National Institute of Biomedical Imaging and Bioengineering, and through generous contributions from the following: AbbVie, Alzheimer’s Association; Alzheimer’s Drug Discovery Foundation; Araclon Biotech; BioClinica, Inc.; Biogen; Bristol-Myers Squibb Company; CereSpir, Inc.; Eisai Inc.; Elan Pharmaceuticals, Inc.; Eli Lilly and Company; EuroImmun; F. Hoffmann-La Roche Ltd and its affiliated company Genentech, Inc.; Fujirebio; GE Healthcare; IXICO Ltd.; Janssen Alzheimer Immunotherapy Research & Development, LLC.; Johnson & Johnson Pharmaceutical Research & Development LLC.; Lumosity; Lundbeck; Merck & Co., Inc.; Meso Scale Diagnostics, LLC.; NeuroRx Research; Neurotrack Technologies; Novartis Pharmaceuticals Corporation; Pfizer Inc.; Piramal Imaging; Servier; Takeda Pharmaceutical Company; and Transition Therapeutics. The Canadian Institutes of Health Research is providing funds to support ADNI clinical sites in Canada. Private sector contributions are facilitated by the Foundation for the National Institutes of Health (www.fnih.org). The grantee organization is the Northern California Institute for Research and Education, and the study is coordinated by the Alzheimer’s Disease Cooperative Study at the University of California, San Diego. ADNI data are disseminated by the Laboratory for Neuro Imaging at the University of Southern California.

## Author contributions

YIM conceived the software’s main interfaces, implemented the analytical source codes (for cTI, ESM, MCM and pTIF), preprocessed and analyzed the presented data, and wrote the manuscript and tutorial drafts. FC implemented the software’s visualization interface *NeuroPM-viewer*. AA, RB and LSR tested early software versions with real data, providing valuable feedback. AK assisted with software compilation to standalone applications for public release. QA and YIM prepared the software’s web page. All authors (except ADNI) contributed to constructive discussions regarding manuscript preparation. ADNI acquired the analyzed data.

## SUPPLEMENTARY INFORMATION (one table)

**Table S1.** Main demographic characteristics for the two populations.

| **Variable** | **HBTRC**<br>**(N=736)** | **ADNI**<br>**(N=911)** |
| --- | --- | --- |
| Women | 354 (47.9%) | 424 (46.5%) |
| Age (years) | 70.8 (15.10) | 72.9 (7.21) |
| Education (years) | - | 16.3 (2.69) |
| HC | 173 (23%) | 266 (29.2%) |
| Diseased (MCI, AD) | 563 (77%) | 645 (70.8%) |

Data are number (%) or mean (std).

